# How well does SARS-CoV-2 spread in hospitals?

**DOI:** 10.1101/2021.09.28.21264066

**Authors:** George Shirreff, Jean-Ralph Zahar, Simon Cauchemez, Laura Temime, Lulla Opatowski, EMEA-MESuRS working group on the nosocomial modelling of SARS-CoV-2

## Abstract

Covid-19 poses significant risk of nosocomial transmission, and preventing this requires good estimates of the basic reproduction number R_0_ in hospitals and care facilities, but these are currently lacking. Such estimates are challenging due to small population sizes in these facilities and inconsistent testing practices.

We estimate the patient-to-patient R_0_ and daily transmission rate of SARS-CoV-2 using data from a closely monitored hospital outbreak in Paris 2020 during the first wave. We use a realistic epidemic model which accounts for progressive stages of infection, stochastic effects and a large proportion of asymptomatic infections. Innovatively, we explicitly include changes in testing capacity over time, as well as the evolving sensitivity of PCR testing at different stages of infection. We conduct rigorous statistical inference using iterative particle filtering to fit the model to the observed patient data and validate this methodology using simulation.

We provide estimates for R_0_ across the entire hospital (2.6) and in individual wards (from 3 to 15), possibly reflecting heterogeneity in contact patterns or control measures. An obligatory mask-wearing policy introduced during the outbreak is likely to have changed the R_0_, and we estimate values before (8.7) and after (1.3) its introduction, corresponding to a policy efficacy of 85%.

## Introduction

The Severe Acute Respiratory Syndrome Coronavirus 2 (SARS-CoV-2) virus has resulted in 136 million confirmed cases and 2.94 million deaths of which 4.99 million and 99,000 respectively have occurred in France ([1] as of 13^th^ April 2021). Despite sweeping control measures, the circulation of SARS-CoV-2 has remained high in many countries, posing a major threat to elderly individuals and individuals with comorbidities, who have poorer clinical outcomes from SARS-CoV-2 infection [2,3]. The protection of fragile individuals necessitates particular vigilance to prevent the spread of SARS-CoV-2 infection within hospitals or long-term care facilities (LTCFs) such as nursing homes. Indeed, nosocomial outbreaks of SARS-CoV-2 have been an important issue since the beginning of the pandemic, with many outbreaks in hospitals or care facilities, often with high attack rates and mortality [4]. More generally, LTCFs are known to be particularly vulnerable to transmission of respiratory pathogens [5].

To control nosocomial SARS-CoV-2 spread, several measures have progressively been implemented in healthcare settings worldwide. These include reinforced contact precautions such as masking, testing strategies among both patients and staff, isolation, visitor restrictions [4], and more recently vaccination [6]. However, the risk of viral transmission and its drivers among hospital patients and staff, and the effectiveness of these control measures, remain incompletely understood, and outbreaks still occur [4,7,8].

The basic reproduction number, R_0_, refers to the number of secondary infections caused by a single index infection in an otherwise susceptible population. This has been widely used as an indicator of SARS-CoV-2 epidemic risk [9], and has also proved valuable for evaluating testing strategies and other preventive measures within healthcare settings [10]. However, very few estimates of reproduction numbers in healthcare settings are available to this day. Due to differences in contact patterns between the general population and within care facilities, the healthcare R_0_ may be expected to both differ from the R_0_ in the general community and vary widely between different types of facilities [11]. Hence, better understanding of SARS-CoV-2 transmission and the impact of control measures in a given care setting first requires a robust estimate of the R_0_ in this specific setting.

However, this is more challenging than estimating the value in the community. The population sizes of institutions are small and therefore epidemics are highly stochastic, with a high risk of extinction. There will usually be more data available from hospitals or services with more cases. Furthermore, testing is rarely conducted at random over time and between patients. Most available data from hospital outbreaks consists of a distribution of positive tests over time, but in a context of evolving testing policy and capacity. At the beginning of the pandemic, in most countries, no standard strategy or recommendation existed on how surveillance should be carried out and tests distributed. They were mostly conducted on symptomatic patients, with possible contact tracing around detected cases. Yet, unreported symptomatic or asymptomatic cases may represent a substantial fraction of the transmissions, with little data on the testing policy itself to estimate how many have fallen through the gaps.

Here, we aimed to better understand the transmission of SARS-CoV-2 in LTCFs and to evaluate the efficacy of barrier measures in reducing it. To that aim, we analysed detailed data from a large hospital outbreak which occurred in the first wave in 2020 using a stochastic transmission model which explicitly accounts for testing policy.

## Methods

### Hospital and patient information

Available data came from a LTCF located in Paris, France. The hospital includes three buildings (A, B, and C) each with four floors (0-3), each of which was considered as a separate ward. For the study period of 61 days the results of all valid PCR tests were available for each patient ID. Patient information also included admission and discharge, ward and ward transfers, as well as presence of symptoms at the time of the first positive test. No additional data were collected from patients other than that for clinical use. All patient data were anonymized and only aggregated data are provided in the published work. All dates are given relative to the date of the first positive test.

### Laboratory testing

All patient samples were taken by nasopharyngeal swab. Motives for testing included the appearance of symptoms characteristic of SARS-CoV-2, having had contact with a positive case, or patient transfer. RNA extraction was performed on a NucliSENS® easyMAG® (Biomerieux, Marcy-l’Etoile,France) device, following strictly the manufacturer’s recommendations. RT-PCR was performed on an ABI QuantStudio 7 (Thermofisher) device, using the commercial RealStar® SARS-CoV-2 RT-PCR Kit 1.0 (Altona Diagnostics, Hamburg, Germany) test. Briefly, 10 µL of RNA is added to the 20 µL RT-PCR mix. Two targets are detected, one specific for beta-coronavirus, and one specific for the SARS-COV-2 strains. Internal Control was added in the lysis buffer to validate both extraction and amplification steps.

### Model description

The spread of infection was modelled using a modified stochastic Susceptible Exposed Infected Recovered (SEIR) model (Figure 1). Equations of the model are provided in Supplementary Methods A.

**Figure 1.**
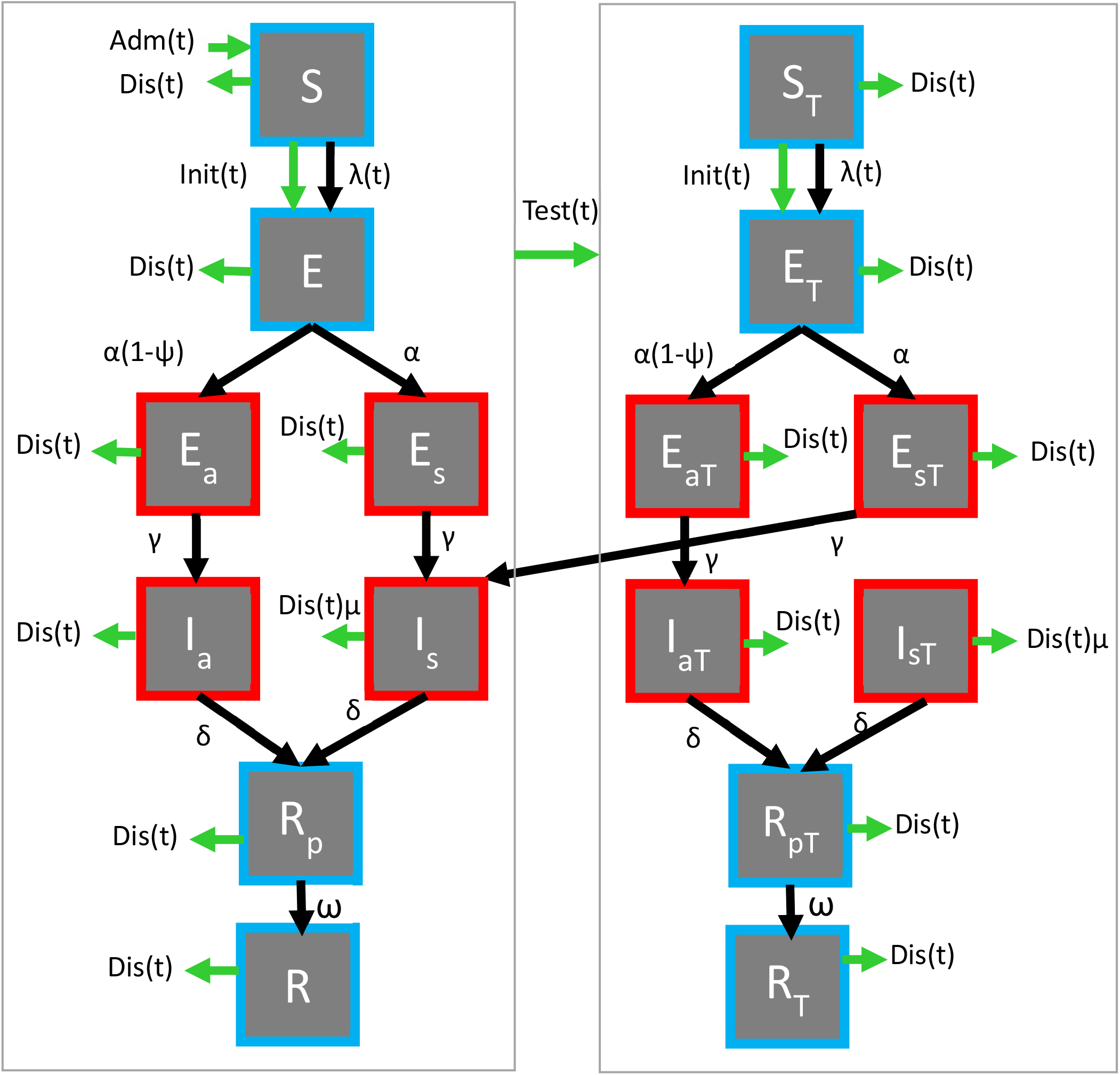
Diagram of the compartmental model, with infectious compartments shown in red and uninfectious in blue. Green arrows refer to processes (Initiation (Init), Admission (Adm), Discharge (Dis) and Testing (Test)) which occur a specified number of times on a given day according to model inputs, whereas black arrow processes correspond to the natural history of infection and are entirely stochastic. The left side shows the trajectory of untested individuals, but if at any point they are tested in state X, they will enter the equivalent tested compartment (X_T_, right panel), which is epidemiologically identical except for the testing rate. The equivalent diagram for the observation process is shown in Supplementary Methods A, Figure 7.

Patients in the susceptible state *S* can become infected by contact with infectious patients. When susceptible individuals become infected they move to the exposed (*E*) compartment for an average duration 1/*α*, during which they have no symptoms and are not yet infectious. Following this, individuals may enter either an asymptomatic or a symptomatic pathway of infection, with the probability of the symptomatic pathway given by *ψ*. Each pathway has an infectious incubation period (*E*_*a*_, *E*_*s*_) before full-blown infection begins (*I*_*a*_, *I*_*s*_), with symptoms appearing only in full-blown infection on the symptomatic pathway (*I*_*s*_). The average duration of the infectious incubation period in the symptomatic pathway is 1/*γ*, and that of full-blown symptomatic infection is 1/*δ*. After full infection, patients recover into a state *R*_*p*_ of average duration 1/*ω* where they are no longer infectious but are still likely to provide a positive test result, before finally entering a fully recovered group (*R*) when the positivity rate diminishes to the false positive rate for uninfected individuals.

The per-capita rate at which susceptible individuals become infected and move to the *E* compartment is led by the force of infection. In brief, the force of infection is proportional to the transmission rate *β* and the number of infected patients, and inversely proportional to the entire population size (details in Supplementary Methods A, Equation 2). At the initiation of the epidemic on date *t*_*init*_, a number (*E*_*init*_) of individuals in non-contagious incubation (*E*) are assumed.

### Transmission rate

The transmission rate of individuals in full-blown symptomatic infection is called *β*. Individuals in the infectious incubation period are assumed to have lower transmission, with infectiousness reduced by a factor *ε*; similarly, infectious individuals in the asymptomatic pathway are also assumed to be less infectious, their transmission rate being reduced by a factor *κ*_*1*_ compared with the symptomatic pathway.

In order to fully understand the outbreak, two distinct assumptions on transmission were modelled and compared. In the primary model, a single transmission rate *β* was assumed to be in effect throughout the period. However, based on knowledge of changing practices within the hospital, we defined a more complex model, called the two-phase model, with each phase characterized by its own transmission rate, *β*_*1*_ and *β*_*2*_, delimited by an inflection date, *t*_*inflect*_. Potential values for this date ranged from day 1 (the date of the first positive sample) to day 16 of the study period (more than a week after the introduction of contact precautions and generalized lockdown in France).

The basic reproduction number can be directly computed for each stage of infection from the transmission rate and duration of each infectious stage as well as the probability of becoming symptomatic (Supplementary Methods A, Equation 4). For the two-phase model, we report an average of the R_0_ in each phase weighted by its duration (Equation 5).

### Observation model

Identification of infected patients was incomplete due to a substantial proportion of asymptomatic infections, imperfect test sensitivity and irregular availability of tests. In order to take into account this imperfect reporting of cases, we added an observation model on top of the transmission model (Supplementary Methods A, Figure 7).

The model tracked the testing status of individuals so that, while any patient could be tested even if they had been previously, retesting occurred at a reduced rate. The relative rate of resting, φ, was estimated directly from the number of tests and retests in the data (Supplementary Methods B). All individuals are initially untested, but upon being tested, they move to an equivalent “tested” state (the tested equivalent of state *X* is *X*_*T*_), where they are less likely to receive a test. The tested status does not change the rates of infectiousness or disease progression, but upon developing symptoms, an individual will lose their tested status and rejoin the untested track (see Figure 1), which allows the model to account for increased testing which occurs when symptoms appear in a patient. The force of infection and initiation of infection occur from the *S* to the *E* compartment, and from the *S*_*T*_ to the *E*_*T*_ compartment, according to the relative size of *S* and *S*_*T*_.

Direct inputs to the model determine the number of admissions, discharges and tests per day, which are given by the data (Supplementary Methods B, Figure 8). Patients are admitted in a susceptible untested state and discharged at random from any state (with a relative rate *μ* for patients with full-blown symptomatic infection). On a given day when tests were performed, if there were individuals who had not been tested since becoming symptomatic, these were tested with priority, and any remaining tests were conducted at random on the rest of the population (Supplementary Methods A, Figure 7).

### Statistical inference

The likelihood was calculated by comparing the number of positive and negative cases on each day with the internal model state via the observation process, assuming a binomial distribution (Supplementary Methods B, Equation 23).

Given the strong stochasticity of the underlying model due to small population size and incomplete observation of the state of each individual, estimating robust likelihoods for given combinations of parameters and estimating the parameter values themselves is challenging. Iterative filtering [18,19] can be used to estimate parameters for a partially observed stochastic process by simultaneously estimating the internal state of the model.

In addition to estimating both transmission rates, (*β*, or *β*_*1*_ and *β*_*2*_), we also estimated the virus introduction time, *t*_*init*_, while *E*_*init*_, the initial number of infections, was fixed to 1. For each analysis (comprising the same model, dataset and fixed parameter values), the confidence intervals (CIs) for the estimated parameters were calculated by comparison with the maximum likelihood. Further details are in Supplementary Methods B.

Model comparison was conducted by calculating the Akaike Information Criterion (AIC). The model and statistical inference methodology were implemented using the *pomp* package [20] in *R* [21]. Further details on the statistical inference are in Supplementary Methods B.

### Model inference validation

As a preliminary step we tested the model and inference methodology on synthetic data to ensure that known simulated transmission rates (*β, or β*_*1*_ and *β*_*2*_) and *t*_*init*_ could be recovered by statistical inference. Several datasets, representing numbers of “observed” positive cases, were generated from model simulations to represent the transmission within the whole hospital and the individual wards over a three month observation period (corresponding to days -39 to +50 relative to the first positive sample), and in each case the real data on numbers of daily tests, admissions, discharges and number of patients were used.

For each known set of parameter values, the median value of the estimated parameters was identified and compared with their known values.

### Analyses at the hospital and ward levels

In the principal analysis the data were analysed at the hospital level assuming homogeneous mixing across all buildings and wards. We censored the data from day 51 onwards, as the containment policy in the hospital began to change after this point.

We also analysed the data at the ward level, in which a separate model and parameter values were estimated for each ward assuming it to be a homogeneous mixing environment. Twenty-three patients were excluded from this analysis because the ward in which they were tested was not available. Again, the data were censored from day 51 onwards.

### Simulated epidemic curves

Following identification of sets of parameter values with likelihoods within the 95% CI relative to maximum likelihood, these sets parameter values were sampled with replacement 1000 times, and each time an epidemic was simulated. Those which went to extinction (having fewer than 3 cumulative infections) were excluded, and the remaining epidemics were used to calculate the median and 95% CI for relevant epidemic variables (number of positive tests, detected and undetected symptomatic and asymptomatic prevalence) for each date. Further details on the estimation of the size of undetected cases is in Supplementary Methods A.

### Sensitivity analysis

Sensitivity analysis was conducted to identify parameters whose variation most affected our estimated parameters. We perturbed the input parameters, usually using the lower and upper bound of the reported CI in the literature, and replicated the analysis.

## Results

A total of 459 patients were present in the hospital during the study period. PCR testing began on day -6 and 312 of these patients subsequently had a definitive PCR test by day 50. The first positive test was recorded on day 1, and 152 patients tested positive by the end of day 50. The distribution of tests in the hospital and in each ward are shown in Figure 2 (left and centre, respectively). The Secondary Attack Rates (SARs) differed substantially between wards (Figure 2, right) ranging from 3 to 50%, while the overall SAR was 33%.

**Figure 2.**
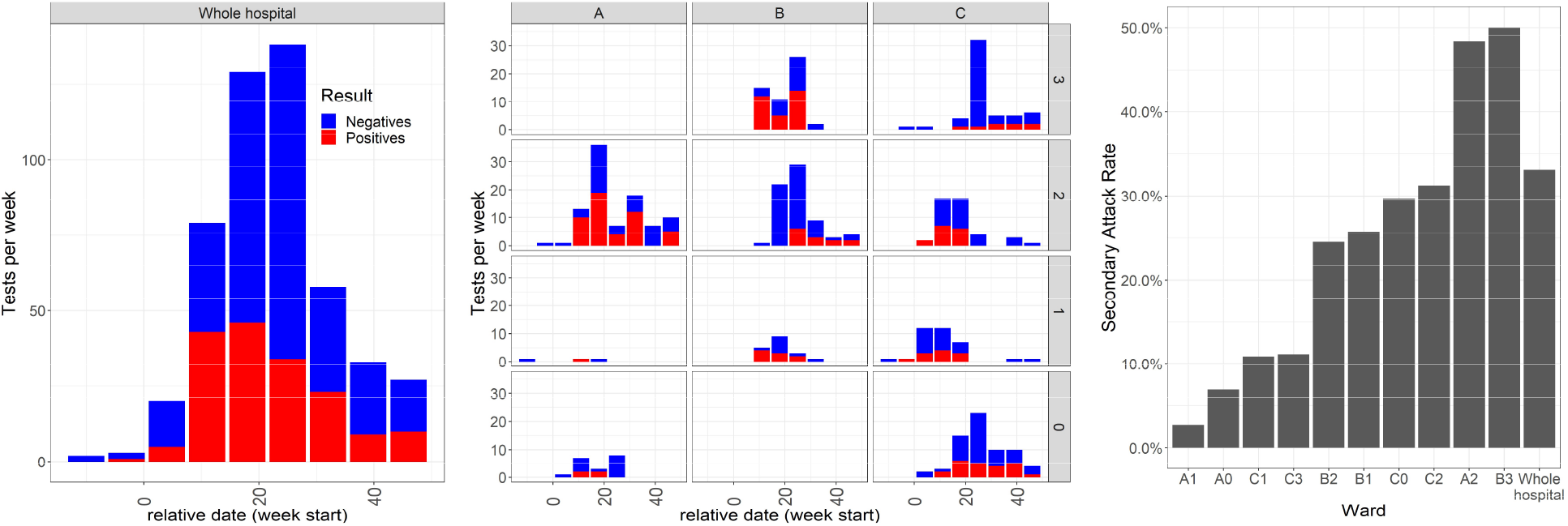
Epidemic curve across all wards of the hospital (left) and in each ward separately (centre), showing the number of tests and their results aggregated by week. Numbers of positive (red) and negative (blue) tests are stacked. Secondary attack rates (SAR) (right) were calculated as the ratio of the number of patients receiving a positive test to the total number of patients present at any time during the period of interest. The letters A-C refer to the buildings and the numbers 0-3 the floors of each building.

### Model inference validation results

The validation of parameter inference for the one-phase and the two-phase model against simulated datasets at the scale of the whole hospital are shown in Supplementary Results A, Figure 10 and Figure 11, while validation for the one-phase model on datasets representing individual wards is shown in Supplementary Results A, Figure 12. These results suggest that there was sufficient power at the level of the whole hospital to recover parameters with relatively good accuracy. At the ward-level, however, this was not always the case, and we restricted our subsequent analysis of wards to only those where the recovered estimates did not deviate excessively. Further details are described in Supplementary Results A.

### Whole hospital analysis

Results from the estimations of transmission rates at the whole hospital level are provided in Table 2. Further details and visual representations on the range of estimates under different assumptions are given in Supplementary Results B.

**Table 1.**
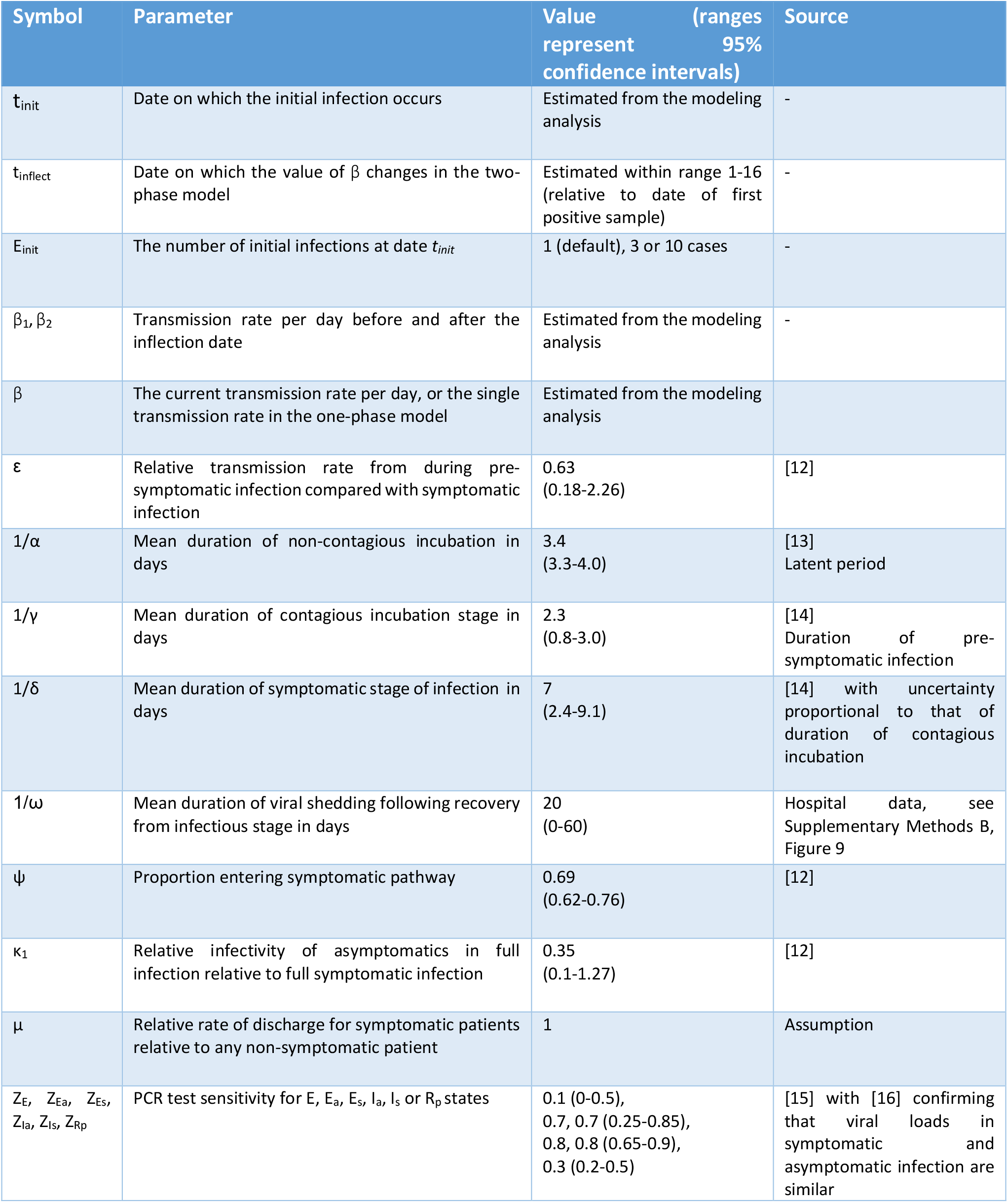
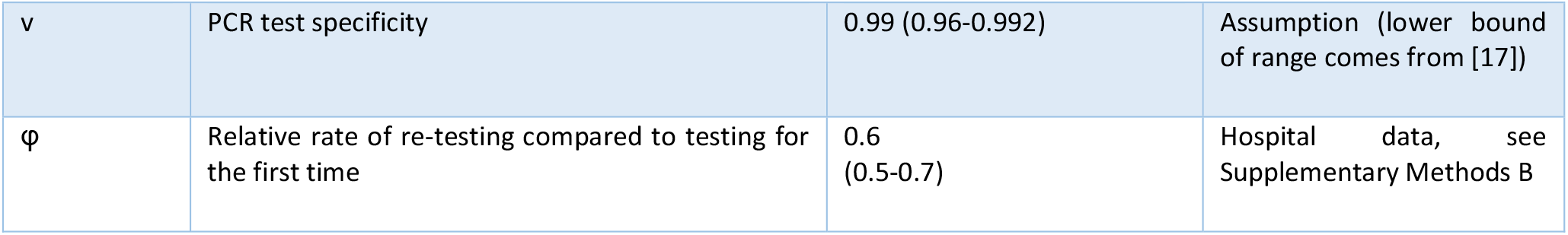
Parameter symbols, descriptions, and assumed values. Additional symbols are described in the methods section and in Supplementary Methods C, Table 4, for reference.

| Symbol | Parameter | Value (ranges represent 95% confidence intervals) | Source |
| --- | --- | --- | --- |
| $t_{init}$ | Date on which the initial infection occurs | Estimated from the modeling analysis | - |
| $t_{inflect}$ | Date on which the value of $\beta$ changes in the two-phase model | Estimated within range 1-16 (relative to date of first positive sample) | - |
| $E_{init}$ | The number of initial infections at date $t_{init}$ | 1 (default), 3 or 10 cases | - |
| $\beta_1, \beta_2$ | Transmission rate per day before and after the inflection date | Estimated from the modeling analysis | - |
| $\beta$ | The current transmission rate per day, or the single transmission rate in the one-phase model | Estimated from the modeling analysis | |
| $\varepsilon$ | Relative transmission rate from during pre-symptomatic infection compared with symptomatic infection | 0.63 (0.18-2.26) | [12] |
| $1/\alpha$ | Mean duration of non-contagious incubation in days | 3.4 (3.3-4.0) | [13]<br>Latent period |
| $1/\gamma$ | Mean duration of contagious incubation stage in days | 2.3 (0.8-3.0) | [14]<br>Duration of pre-symptomatic infection |
| $1/\delta$ | Mean duration of symptomatic stage of infection in days | 7 (2.4-9.1) | [14] with uncertainty proportional to that of duration of contagious incubation |
| $1/\omega$ | Mean duration of viral shedding following recovery from infectious stage in days | 20 (0-60) | Hospital data, see Supplementary Methods B, Figure 9 |
| $\psi$ | Proportion entering symptomatic pathway | 0.69 (0.62-0.76) | [12] |
| $\kappa_1$ | Relative infectivity of asymptomatics in full infection relative to full symptomatic infection | 0.35 (0.1-1.27) | [12] |
| $\mu$ | Relative rate of discharge for symptomatic patients relative to any non-symptomatic patient | 1 | Assumption |
| $Z_E, Z_{Ea}, Z_{Es}, Z_{Ia}, Z_{Is}, Z_{Rp}$ | PCR test sensitivity for E, $E_a$, $E_s$, $I_a$, $I_s$ or $R_p$ states | 0.1 (0-0.5), 0.7, 0.7 (0.25-0.85), 0.8, 0.8 (0.65-0.9), 0.3 (0.2-0.5) | [15] with [16] confirming that viral loads in symptomatic and asymptomatic infection are similar |
| $v$ | PCR test specificity | 0.99 (0.96-0.992) | Assumption (lower bound of range comes from [17]) |
| $\phi$ | Relative rate of re-testing compared to testing for the first time | 0.6 (0.5-0.7) | Hospital data, see Supplementary Methods B |

**Table 2.** Best estimates and their ranges for β, R_0_ and t_init_ (in the case of the one-phase model) and β_1_, β_2_, R_0_ in each phase and combined, and t_init_ (in the two-phase model). The values of E_init_ and t_inflect_ were fixed at 1 and day 12 respectively. The R_0_ values were calculated using Equations 4 and 5. The intervention efficacy was calculated as 1-β_2_/β_1_. Days for t_init_ are relative to the first positive sample.

| Model | $\beta$ | | | | $R_0$ | | $t_{init}$ | AIC |
| --- | --- | --- | --- | --- | --- | --- | --- | --- |
| One-phase | 0.38<br>(0.30-0.60) | - | - | - | 2.6<br>(2.0- 4.1) | - | -22 (-39 – -4) | 657.33 |
| Model | $\beta_1$ | $\beta_2$ | $R_0$ before | $R_0$ after | $R_0$ combined | Intervention efficacy | $t_{init}$ | AIC |
| Two-phase | 1.28<br>(0.76-2.40) | 0.19<br>(0.10-0.30) | 8.72<br>(5.14-16.32) | 1.33<br>(0.68-2.04) | 5.72<br>(3.62- 8.70) | 0.85<br>(0.66-0.94) | -4 (-25 – -1) | 628.85 |

In the two-phase model, using day 12 as *t*_*inflect*_ gave the best model fit (Supplementary Table 6). This more complex model (Table 2 and Supplementary Table 7) proved a better fit to the data than the one-phase model, as measured by the AIC. The two-phase model also visually better captured the early peak in cases since all data points fell within the 95% range of simulations (Figure 3). A more direct comparison of the proximity of the simulations to the data for each model can be seen in Supplementary Figure 18.

**Figure 3.**
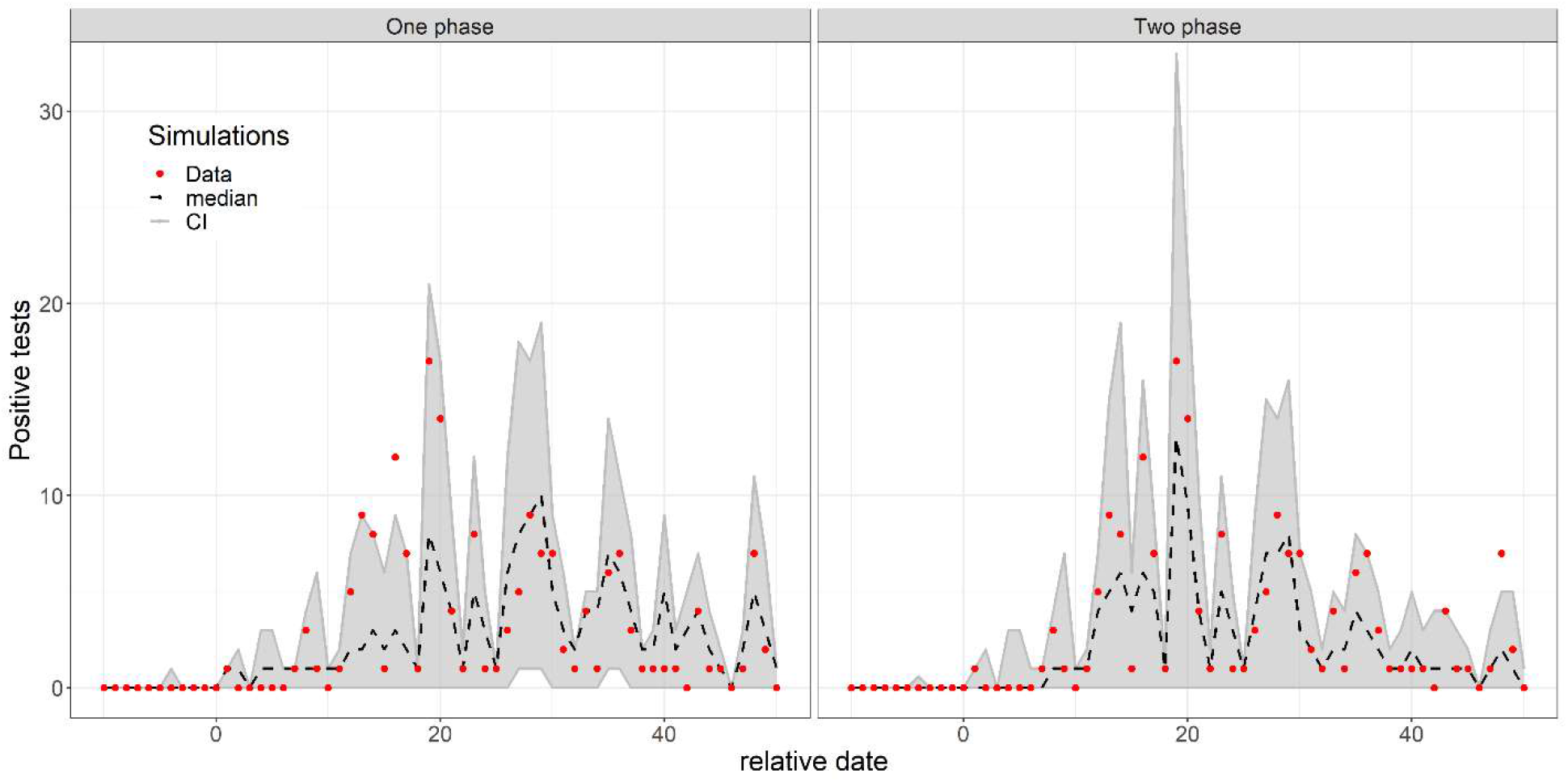
Simulations using the set of parameters with likelihoods which fall within the 95% CI relative to the maximum likelihood, for the one-phase model (left) and the two-phase model (right). The exact parameter combinations used in each, correspond to the values above the dotted line in the scatterplots in Supplementary Figure 14 and Figure 16 where E_init_ = 1. The red points show the observed number of positive tests in the data, the grey area the range in which 95% of the simulation values fall, and the black dashed line the median across that date for all simulations. Extinct epidemics (having fewer than 3 cumulative cases) were excluded from the distribution.

Interestingly, in the best model, a notable difference was observed between the transmission rates estimated before and after day 12, which we assume to be the date of effective contact precaution implementation. In the first phase the rate was estimated at 1.3 (95%CI 0.8-2.4) infections per patient per day in symptomatic infection, corresponding to an R_0_ of 8.7 (95%CI 5.1-16.3), and in the second phase the rate was 0.19 (95%CI 0.10-0.30) corresponding to an R_0_ of 1.3 (0.7-2.0). This translates to a decrease of the transmission risk by 85% (95% CI 66-94%) in the second phase after the generalised implementation of contact precautions in the hospital. The range of likely parameter values are shown in more detail in the scatterplots in Supplementary Figure 14 and Figure 16. The value of *t*_*inflect*_ was selected for how well it explained the data, and the chosen date occurs shortly after the introduction of an obligatory mask-wearing policy and cancellation of all group activities between patients (day 6). Although the value of *t*_*inflect*_ had a substantial effect on the absolute values of the transmission rates, the size of the decrease in transmission rate was relatively stable (81% – 89%, Supplementary Table 6).

### Individual wards analysis

Table 3 provides estimates obtained for each individual ward for which the model could be validated, using the one-phase model. Results for the ward-level using the two-phase model, and the comparison between the two models are shown in Supplementary Results C, Table 8, although the one-phase model outperforms the two-phase in three out of four wards.

**Table 3.** Ward characteristics and parameter estimates. Estimates and their 95% CI for β, R_0_ and t_init_ from the fitting of the one-phase model to data from each ward (E_init_=1). In many instances, the upper bound of the CI for β, and in some the most likely value of β as well, could not be estimated due to a flat likelihood surface, in which case the value is given as N.E.=not estimated. The R_0_ values were calculated using Equation 4.

| Ward | Beds | Cumulative patients | Day of first positive | Cases | $\beta$ | $R_0$ | $t_{init}$ |
| --- | --- | --- | --- | --- | --- | --- | --- |
| A2 | 48 | 62 | 11 | 30 | 1.29<br>(0.51- N.E.) | 8.76<br>(3.47- N.E.) | 2 (-14 – 29) |
| C0 | 37 | 74 | 16 | 22 | 0.56<br>(0.22- N.E.) | 3.79<br>(1.50- N.E.) | 4 (-39 – 9) |
| C2 | 37 | 48 | 7 | 15 | 2.13<br>(0.29- N.E.) | 14.46<br>(1.97- N.E.) | -8 (-39 – -14) |
| C3 | 37 | 63 | 24 | 7 | 0.42<br>(0.11-1.30) | 2.87<br>(0.75- 8.84) | 19 (-9 – 21) |

Point estimates for *β* ranged from 0.42 to 2.13 across the studied wards. Only in one ward, C3, was it possible to calculate an upper bound for the transmission rate, and the resulting range estimate of 0.42 (0.11-1.30) corresponds to an R_0_ of 2.87 (0.75-8.84). However, a lower bound was estimable in each case, with the highest value 0.51 in ward A2 corresponding to a minimum R_0_ of 3.47. The observed data consistently fell within or on the boundary of the 95% confidence range of simulations (Figure 4).

**Figure 4.**
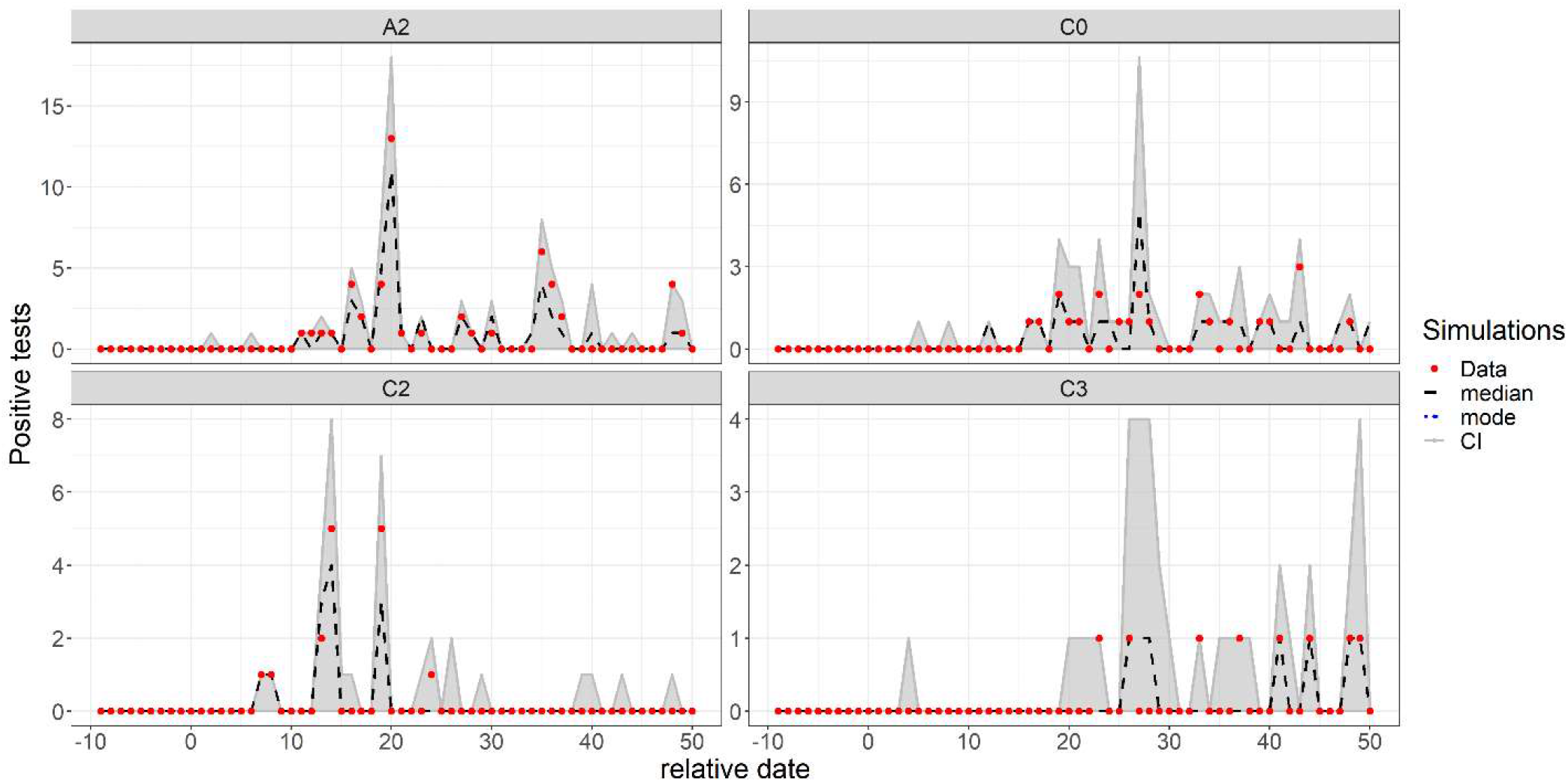
Simulations of the one-phase model using the sets of parameters with likelihoods which fall within the 95% CI of the maximum likelihood (as shown in Supplementary Results B, Figure 19). The red points show the observed number of positive tests in the data, the grey area the range in which 95% of the simulation values fall, and the black dashed line the median across that date for all simulations. Extinct epidemics (having fewer than 3 cumulative cases) were excluded from the distribution.

### Undetected epidemic dynamics

As testing is incomplete for all infections, and many infections are asymptomatic and therefore less likely to be tested, the prevalence of undetected infections is large. Using simulated epidemics with likely values of the relevant parameters (as in Figure 3), we are able to show the scale of the detected and undetected cases in the hospital (Figure 5). At peak prevalence of infectious individuals, the proportion undetected was estimated at 60%, and approached 30% by the end of the period of interest. The median cumulative incidence estimated by the model was 199 patients, and when compared with 152 observed positive patients suggests approximately 25% of cases remained ultimately undetected.

**Figure 5.**
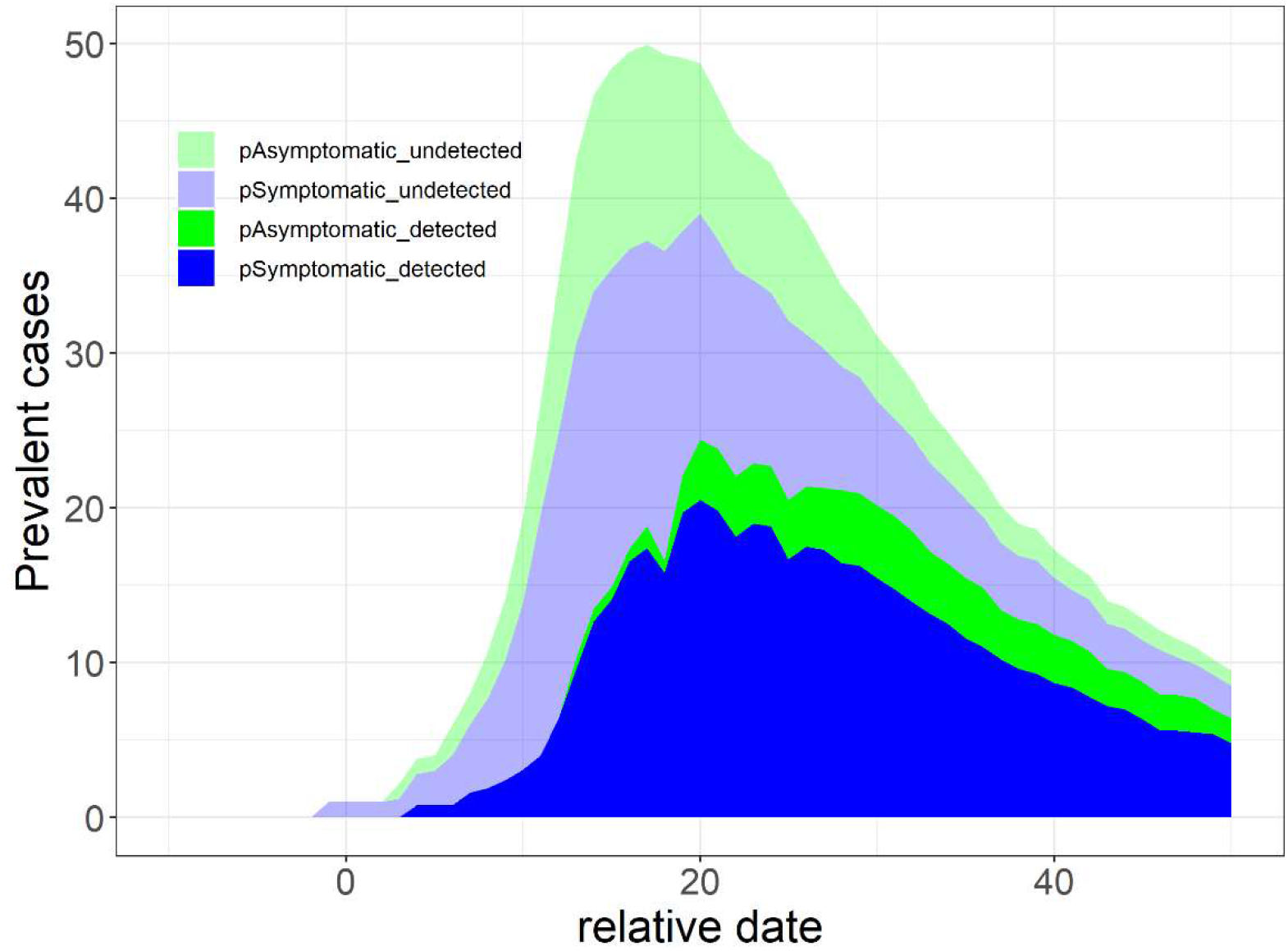
Stacked prevalence of detected and undetected symptomatic and asymptomatic infections in simulated epidemics at the scale of the whole hospital, using the set of parameters with likelihoods which fall within the 95% CI relative to the maximum likelihood using the two-phase model. After exclusion of extinct simulations (having fewer than 3 cumulative cases), the median of each prevalence measure was calculated for each date.

**Figure 6.**
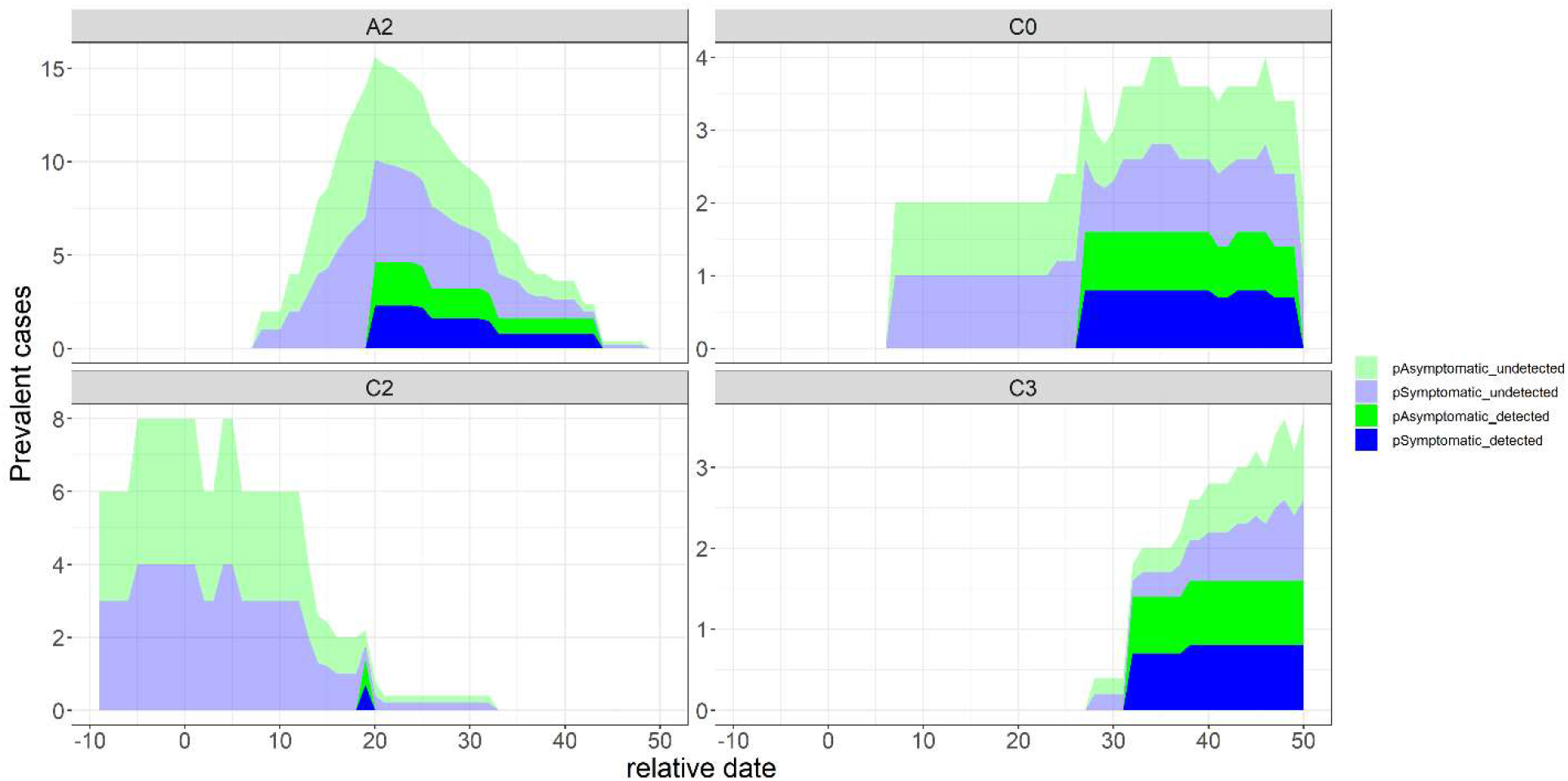
Stacked prevalence of detected and undetected symptomatic and asymptomatic infections in simulated epidemics at the scale of individual wards, using the set of parameters with likelihoods which fall within the 95% CI relative to the maximum likelihood using the one-phase model. After exclusion of extinct simulations (having fewer than 3 cumulative cases), the median of each prevalence measure was calculated for each date.

### Sensitivity analysis results

The perturbation of most parameters had relatively minor effects on the estimated transmission rates for the two phases, as well as on *t*_*init*_ (Supplementary Results D, Figure 20). The transmission rate in the second phase, *β*_*2*_, was the most sensitive, and most markedly to the duration of symptomatic infection (1/δ).

## Discussion

In this paper we analysed data from a LTCF outbreak using a transmission model of patient-to-patient infection with SARS-CoV-2 to estimate transmission rates, either across the entire hospital or in individual wards, and over one or two phases of transmission delimited by a specific change date (*t*_*inflect*_) corresponding to the implementation of preventive measures.

Validation of the parameter inference for the two-phase model on synthetic data demonstrated that we were able to estimate two separate transmission rates occurring either side of *t*_*inflect*_, and therefore that we could estimate the effectiveness of the measures implemented for the second phase after generalized awareness of the presence of the outbreak. We found that the two-phase model was better supported by the data aggregated across the entire hospital than a model with a single transmission rate, and it better captured the early peak in cases. The early phase transmission rate (1.3 transmissions per patient per day) and late phase rate (0.19) correspond to early and late R_0_ values of 8.7 and 1.3, respectively. This change in transmission rate can largely be explained by the initial absence of preventive measures and the delay in their introduction, followed by the recommendation of contact precautions on day 6 and its gradual implementation over the following week. Under this assumption, the measures introduced were 85% (66-94%) effective at reducing transmission.

Studies on other respiratory patterns have quantified the protective effect in healthcare of mask wearing by patients [22] and by healthcare workers (HCWs) [23], the latter showing contact precautions had a protective efficacy for both patients and personnel of around 85%. A meta-analysis of studies of mask use suggested that in healthcare settings, face masks had 67% protective efficacy against SARS, and that N95 respirators had 96% efficacy against SARS, MERS and SARS-CoV-2 [24]. However, the one study examining the effect of contact precautions against SARS-CoV-2 only examined a protective effect for HCWs, which was unquantifiable as there were no infections in the masked group [25]. Several modelling studies have attempted to quantify the level of mask wearing which would prevent epidemic spread of SARS-CoV-2 in the community [26–29], but none have done so in healthcare environments. To our knowledge no study has used a detailed epidemic model to quantify the effect of a mask-wearing policy at protecting patients in healthcare.

Our estimate for the transmission rate (1.3, 0.8-2.4) and reproduction number (8.7, 5.1-16.3) during the first phase are high, suggesting an explosive outbreak, which is consistent with our high SAR (33%) in a 350-bed facility. Our estimate of the transmission rate (0.19, 0.10-0.30) and reproduction number (1.3, 0.7-2.0) in the second phase following the updated policy may be more representative of current transmission rates in hospitals which are able to provide and encourage the use of such personal protective equipment.

To our knowledge, only three other studies have published estimates of reproduction numbers in healthcare. Tang et al. [30] used the cumulative proportion of polymerase chain reaction (PCR) positives in the exponential growth phase of a new epidemic as the attack rate, from which an expedient estimate of R_0_ was calculated for both patients (1.13) and hospital staff (1.21), out of 7840 and 523 tested, and SAR of 21% and 29% respectively. The study assumed a single index case, which if incorrect would imply an overestimate of the attack rate [31]. However, as testing was only systematically performed on symptomatic individuals, any additional contribution of asymptomatic infections may lead to underestimation. While this is an appropriate use of the available data, this study was also not able to provide a range for the R_0_ estimates. Reyné et al. also estimated a basic reproduction number of 1.021 with 95% CI of 1.018-1.024 across 12 nursing homes based on a single introduction per floor of each institution and a SAR of 4.1% out of 930 residents [32]. The heterogeneity of transmission between different services was also demonstrated in a review and meta-analysis by Thompson et al. [33] which examined four different studies in healthcare settings to calculate the observed reproduction number (R_obs_), defined as the average number of secondary cases of SARS-CoV-2 per index case. While R_obs_ is a measure of epidemic potential, it assumes that there is only a single generation of infection, and if this assumption is incorrect then it will imply an overestimate of the effective reproduction number. They calculated an average of 1.18, but with much heterogeneity between studies (one of 4.5, and three less than 0.25).

In order to assess to which extent the estimates at the whole hospital level may vary when looking at smaller scales, we attempted to fit a model to data from each ward. At the ward level we assumed a single phase of transmission and a single index case. Nevertheless, estimating the upper bounds of the transmission rates proved challenging, probably due to strong stochasticity in small populations and scarcity of observed cases, which is an inherent feature of SARS-CoV-2 where a significant proportion of infected individuals stay asymptomatic. However, our validation analyses suggested that point estimates for transmission rates across the wards could be consistently estimated. Applied to our dataset, estimated values ranged widely, from 0.4 to 2.1 per patient per day, corresponding to R_0_ values from 2.9 to 14.5 depending on the ward. This heterogeneity may have been driven by differences in the application of preventive measures, both in terms of timing and of observance. Contact patterns between staff and patients or among patients may also differ between wards according to their function.

As mentioned earlier, a major difficulty in analyzing and estimating parameters for hospital outbreaks or more generally closed outbreaks of SARS-CoV-2 in small populations lies in the consideration of testing practice. Because tests are not carried out systematically, reported incidence of cases is imperfect and an important part of the process is unobserved. Indeed, here the reporting of infected cases over the study period relied on the results of positive tests. First, as already mentioned, a substantial proportion of infectious individuals were not symptomatic, therefore possibly not detected nor tested. Second, PCR test sensitivity is not perfect and depends on the timing since infection. We addressed the issues around imperfect observation by using a detailed and realistic observation process, which also allows us to quantify the burden of undetected symptomatic and asymptomatic infections. Lastly, testing procedure was not regular, and may have been affected by a large number of factors which are irrelevant to general epidemiological analysis, such as the day of the week, the available testing capacity, or changing strategies at the local scale. We addressed this by using the number of tests per day directly as described in the data, rather than determining the number of tests from the epidemiological state, such as the number of infected individuals. The model also tracked testing status in order to include realistic probabilities for testing and retesting of patients. Although we have not attempted it in this study, the use of this feature to capture tests which arise from contact tracing could allow explicit exploration of testing strategies, especially when combined with a model correlating traced contact and transmission probability [34].

There are several limitations to the analysis as a result of simplifying assumptions. Firstly, when running the entire hospital analysis, the ward structure was not accounted for, even though it could be assumed that the rate of intra-ward transmission is higher than inter-ward. Secondly, we did not take into account the possibility of imported infections other than from the index case(s), preferring to assume that after the very early stages of the outbreak, the force of infection from other patients would substantially outweigh those arriving from the community.

Thirdly, we focused on patients and did not explicitly model acquisition nor transmission by or from HCWs. No data on infection status for the health care workers within the hospital over the study period was available. Health care workers were therefore indirectly modelled as vectors of transmission through the patient-patient infection process. Rates of transmission to HCWs exposed to infectious patients are relatively low [35,36], as well as from HCWs to patients [37], although this may have been less true in the early stages of the pandemic, given low levels of hand hygiene [38]. Ignoring the contribution of HCWs to new infections in the analysis suggests that we may have overestimated the transmission risk from infectious patients. However, our estimates can still be interpreted as valid measures of the nosocomial risk to patients.

Fourthly, we note that the decision to analyze data from this hospital is partly due to the size of the outbreak, implying a selection bias towards a higher transmission rate than would be typical across all hospitals. However, given over 44,000 nosocomial infections reported in France up to 14^th^ February 2021 [39], the vast majority of which consisted of clusters of cases, these results can be interpreted as plausible for a hospital at risk of an outbreak.

Very detailed longitudinal data were available here, including positive and negative tests, as well as admissions, discharges and patients present each day. This analysis could be easily repeated on other similar datasets; the modelling framework could also be easily adapted to include other epidemiological features such as isolation of patients who have tested positive.

## Conclusion

Our results underline both the large potential effect of protective interventions introduced in healthcare settings, and the considerable heterogeneity in transmission rates between hospital wards. In addition, the modelling and statistical inference methodology we developed is straightforward to adapt to other datasets and epidemiological scenarios.

## Supporting information

Supplementary Results

## Data Availability

All data analysed for this study at the whole hospital and ward levels are available from the corresponding author upon request.
The R scripts used to conduct the analysis are available from the corresponding author upon request.

## Abbreviations

AIC: Akaike information criterion
CI: Confidence interval
LTCF: Long-term care facility
PCR: Polymerase chain reaction
POMP: Partially observed Markov process
SAR: Secondary attack rate
SARS-CoV-2: Severe Acute Respiratory Syndrome coronavirus 2
SEIR: Susceptible Exposed Infected Recovered
HCW: Healthcare worker

## Declarations

### Availability of data and material

All data analysed for this study at the whole hospital and ward levels are available from the corresponding author upon request.

### Code availability

The R scripts used to conduct the analysis are available from the corresponding author upon request.

### Author contributions

GS constructed the model, conducted the analysis, and produced the draft and graphics. JRZ produced the data and provided the medical perspective to inform model assumptions. SC made substantial contributions to the interpretation of results. LT and LO conceived the study, had regular input on analysis and interpretation, and contributed to the writing. All authors read, provided comments on and approved the final manuscript.

### Ethics approval

The Comité Local d’Ethique pour la Recherche Clinique des HUPSSD Avicenne-Jean Verdier-René Muret has approved the study as protocol CLEA-2021-190.

### Consent to participate and publish

No additional data were collected from patients other than that for clinical use. All patient data were anonymized and only aggregated data are provided in the published work.

## Acknowledgements

The work was supported directly by internal resources from the French National Institute for Health and Medical Research, the Institut Pasteur, the Conservatoire National des Arts et Métiers, and the University of Versailles-Saint-Quentin-en-Yvelines/University of Paris-Saclay. This study received funding through the MODCOV project from the Fondation de France grant 106059 as part of the alliance framework “Tous unis contre le virus”, the Université Paris-Saclay (AAP Covid-19 2020) and the French government through its National Research Agency project SPHINX-17-CE36-0008-01.

The authors would like to acknowledge the help of the EMEA-MESuRS working group on the nosocomial modelling of SARS-CoV-2 (Audrey Duval, Kévin Jean, Sofía Jijón, Ajmal Oodally, Lulla Opatowski, George Shirreff, David RM Smith, Laura Temime), Niels Hendrickx, Sandrine Jacques and Matthieu Domenech de Cellès.

