## Supplementary Methods for "How well does SARS-CoV-2 spread in hospitals?"

#### A - Mathematical model

##### Observation model

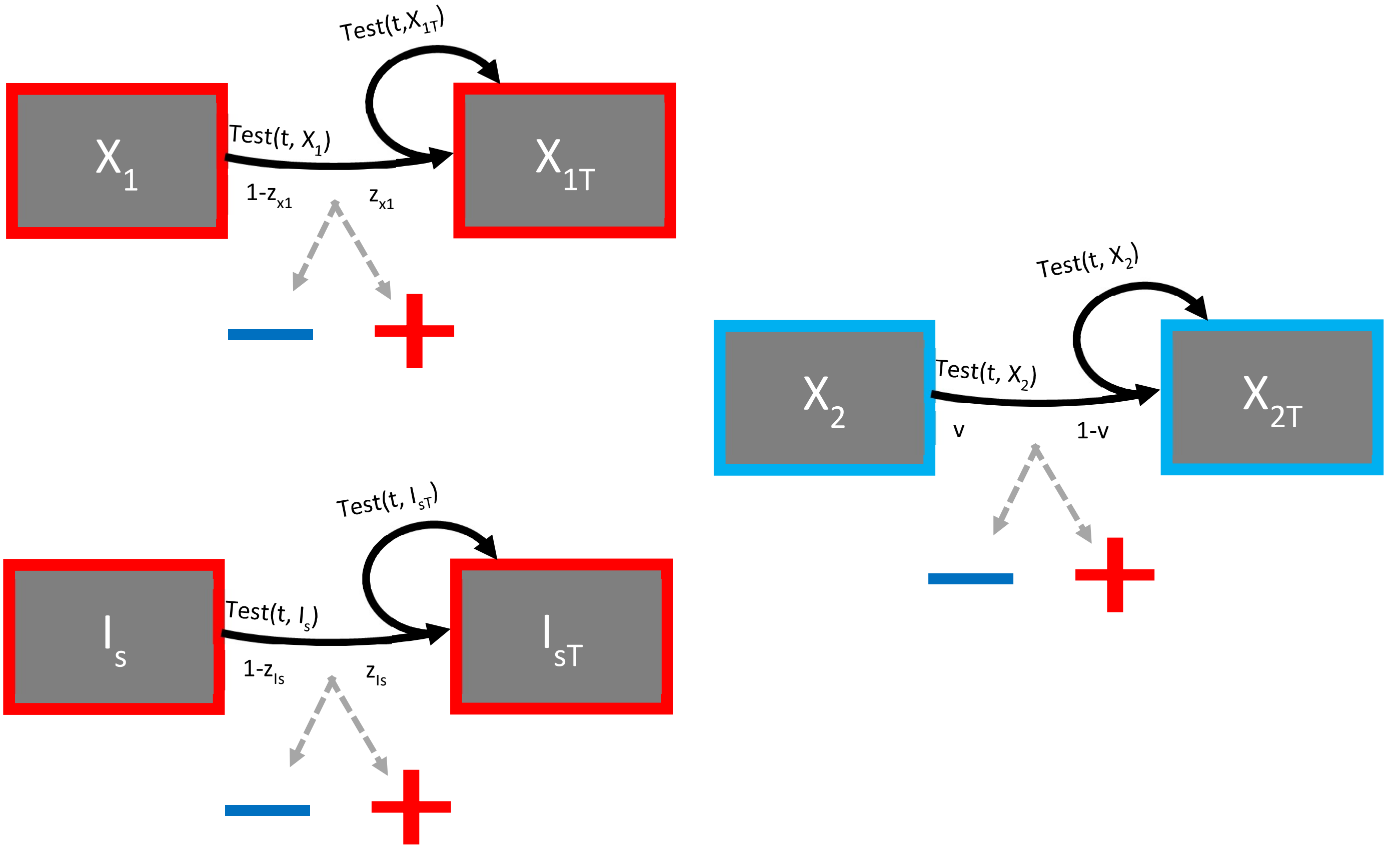


*Figure 7. Illustration of the observation process, where $X_1$ represents any compartment of untested individuals who are shedding virus ($E$, $E_a$, $E_s$, $I_a$, $R_p$) who test positive at their compartment-specific sensitivity rate ($z_{X1}$), and $X_2$ represents any compartment of not recently tested individuals who are not shedding virus (S, R) who test negative at rate v. $X_{1T}$, $I_{sT}$ and $X_{2T}$ and represent their tested counterparts. The symptomatic individuals ($I_s$ and $I_{sT}$) are shown separately because testing is conducted first on the non-recently tested symptomatic group, but retesting is equally likely for symptomatic individuals as non-symptomatic. Upon testing or retesting, the dotted arrows indicate the probabilities of the possible observed outcomes, positive and negative.*

##### Differential equations

The structure of the transmission model shown in Figure 1 and the observation model in Figure 7 can be represented using differential equations which describe the change in state of each compartment in each time step (Equations 1), in which $\lambda(t)$ is the force of infection (Equation 2).

$$\begin{aligned}
\frac{dS}{dt} &= \mathbf{Admission}(\boldsymbol{t}) - \mathbf{Initiation}(\boldsymbol{S},\boldsymbol{t}) - \lambda(t)S - \mathbf{Discharge}(\boldsymbol{S},\boldsymbol{t}) - \mathrm{Test}(\boldsymbol{t},\boldsymbol{S}) \\
\frac{dS_T}{dt} &= -\mathbf{Initiation}(\boldsymbol{t},\boldsymbol{S_T}) - \lambda(t)S_T - \mathbf{Discharge}(\boldsymbol{t},\boldsymbol{S_T}) + \mathrm{Test}(\boldsymbol{t},\boldsymbol{S}) \\
\frac{dE}{dt} &= \mathbf{Initiation}(\boldsymbol{t},\boldsymbol{S}) + \lambda(t)S - \alpha E - \mathbf{Discharge}(\boldsymbol{t},\boldsymbol{E}) - \mathrm{Test}(\boldsymbol{t},\boldsymbol{E})
\end{aligned} \qquad 1$$

$$\frac{dE_T}{dt} = \textbf{Initiation}(\boldsymbol{t}, \boldsymbol{S_T}) + \lambda(t)S_T - \alpha E_T - \textbf{Discharge}(\boldsymbol{t}, \boldsymbol{E_T}) + \textbf{Test}(\boldsymbol{t}, \boldsymbol{E})$$
$$\frac{dE_a}{dt} = \alpha E(1-\psi) - E_a\gamma\kappa_2 - \textbf{Discharge}(\boldsymbol{t}, \boldsymbol{E_a}) - \textbf{Test}(\boldsymbol{t}, \boldsymbol{E_a})$$
$$\frac{dE_{aT}}{dt} = \alpha E_T(1-\psi) - E_{aT}\gamma\kappa_2 - \textbf{Discharge}(\boldsymbol{t}, \boldsymbol{E_{aT}}) + \textbf{Test}(\boldsymbol{t}, \boldsymbol{E_a})$$
$$\frac{dE_s}{dt} = \alpha E\psi - E_s\gamma - \textbf{Discharge}(\boldsymbol{t}, \boldsymbol{E_s}) - \textbf{Test}(\boldsymbol{t}, \boldsymbol{E_s})$$
$$\frac{dE_{sT}}{dt} = \alpha E_T\psi - E_{sT}\gamma - \textbf{Discharge}(\boldsymbol{t}, \boldsymbol{E_{sT}}) + \textbf{Test}(\boldsymbol{t}, \boldsymbol{E_s})$$
$$\frac{dI_a}{dt} = E_a\gamma\kappa_2 - I_a\delta\kappa_3 - \textbf{Discharge}(\boldsymbol{t}, \boldsymbol{I_a}) - \textbf{Test}(\boldsymbol{t}, \boldsymbol{I_a})$$
$$\frac{dI_{aT}}{dt} = E_{aT}\gamma\kappa_2 - I_{aT}\delta\kappa_3 - \textbf{Discharge}(\boldsymbol{t}, \boldsymbol{I_{aT}}) + \textbf{Test}(\boldsymbol{t}, \boldsymbol{I_a})$$
$$\frac{dI_s}{dt} = E_{sT}\gamma + E_s\gamma - I_s\delta - \textbf{Discharge}(\boldsymbol{t}, \boldsymbol{I_s}) - \textbf{Test}(\boldsymbol{t}, \boldsymbol{I_s})$$
$$\frac{dI_{sT}}{dt} = -I_{sT}\delta - \textbf{Discharge}(\boldsymbol{t}, \boldsymbol{I_{sT}}) + \textbf{Test}(\boldsymbol{t}, \boldsymbol{I_s})$$
$$\frac{dR_p}{dt} = I_s\delta + I_a\delta\kappa_3 - \omega R_p - \textbf{Discharge}(\boldsymbol{t}, \boldsymbol{R_p}) - \textbf{Test}(\boldsymbol{t}, \boldsymbol{R_p})$$
$$\frac{dR_{pT}}{dt} = I_{sT}\delta + I_{aT}\delta\kappa_3 - \omega R_{pT} - \textbf{Discharge}(\boldsymbol{t}, \boldsymbol{R_{pT}}) + \textbf{Test}(\boldsymbol{t}, \boldsymbol{R_p})$$
$$\frac{dR}{dt} = \omega R_p - \textbf{Discharge}(\boldsymbol{t}, \boldsymbol{R}) - \textbf{Test}(\boldsymbol{t}, \boldsymbol{R})$$
$$\frac{dR_T}{dt} = \omega R_{pT} - \textbf{Discharge}(\boldsymbol{t}, \boldsymbol{R_T}) + \textbf{Test}(\boldsymbol{t}, \boldsymbol{R})$$

#### Force of infection and $R_0$ calculation

The force of infection acting on susceptible patients (Equation 2) is defined by the infectious populations, the transmission rate *β* and the total population size *N* (Equation 3).

$$\lambda(t) = \frac{\beta\left(I_s + I_{sT} + \varepsilon(E_s + E_{sT}) + \kappa_1(I_a + I_{aT}) + \varepsilon\kappa_1(E_a + E_{aT})\right)}{N} \qquad 2$$

$$N = S + S_T + E + E_T + E_a + E_{aT} + E_s + E_{sT} + I_a + I_{aT} + I_s + I_{sT} + R_p + R_{pT} + R + R_T \qquad 3$$

The value of $R_0$ can be calculated directly from the parameters according to Equation 4, which takes into account the full-blown symptomatic transmission rate *β*, the probability of entering the symptomatic or asymptomatic pathway (*ψ* or 1-*ψ*), the relative transmission rate of each stage of infection (*ε*, $\kappa_1$), and the rate of leaving each stage (γ and δ). In the two-phase model, two values of $R_0$ ($R_{0\ \text{before}}$ and $R_{0\ \text{after}}$) were calculated independently for each phase based on the different transmission rates $\beta_1$ and $\beta_2$ using Equation 4, while a combined value of $R_0$ was calculated as an average weighted by the duration of each phase using Equation 5, with the final date being the end of the study period, day 50.

$$R_0 = \beta\left(\psi\left(\frac{\varepsilon}{\gamma} + \frac{1}{\delta}\right) + (1-\psi)\kappa_1\left(\frac{\varepsilon}{\gamma} + \frac{1}{\delta}\right)\right) \qquad 4$$

$$R_{0\ \text{combined}} = R_{0\ \text{before}}\left(t_{inflect} - t_{init}\right) + R_{0\ \text{after}}\left(\text{final\_date} - t_{inflect}\right) \qquad 5$$

#### Deterministic processes

The bold terms in Equations 1 above refer to flows which are determined in part by the available data from a given day (number of admissions *A*(*d*), of discharges *D*(*d*) and of tests *T*(*d*) as shown in weekly aggregate in Figure 8) or by specific parameter values (SARS-CoV-2 introduction date $t_{init}$ and size $E_{init}$, which together determine the number of daily new infectees *C*(*d*)) in the case of epidemic initiation. This is to ensure that admissions, discharges and number of tests occur each with the same frequency in the model as they do in the data, and that the timing and size of the start of the epidemic is determined by parameter values.

##### Admission

Admissions only occur into the susceptible untested (S) group, and so on a given day *d* the number of admissions into this group is exactly determined by the number of admissions on that day, *A*(*d*):

$$\int_{t=d}^{t=d+1} \mathbf{Admission}(t)dt = A(d) \qquad 6$$

##### Initiation

The expectation of the number of initial infectees from each susceptible group ($S$, $S_T$) is determined by Equations 7 and 8, where *C*(*d*) is equal to $E_{init}$ on day $t_{init}$, and otherwise zero, with the total in both groups being equal to *C*(*d*) (Equation 10):

$$E\big(\mathbf{Initiation}(d,S)\big) = C(d)\frac{S}{S+S_T} \qquad 7$$

$$E\big(\mathbf{Initiation}(d,S_T)\big) = C(d)\frac{S_T}{S+S_T} \qquad 8$$

For simplification, we denote the number of infections initiated on day *d* in compartment *X* by Equation 9, which is equal to the integral of all initiations in all (both) compartments across the whole of the day. The total initiations across all compartments is equal to *C*(*d*) (Equation 10).

$$\mathbf{Initiation}(d,X) = \int_{t=d}^{t=d+1} \mathbf{Initiation}(t,X)dt \qquad 9$$

$$\sum\nolimits_{X\in S,S_T} \mathbf{Initiation}(d,X) = C(d) \qquad 10$$

##### Discharge

The expectation of the number of discharges for a compartment *X* on each day (Equation 11) has different values for symptomatically infected patients ($I_s$ and $I_{sT}$), who have a discharge rate modified by the parameter $\mu$. The denominator of the expectation, *W*, is the total dischargeable population, adjusting for these differences in rate (Equation 12).

$$E\big(\mathbf{Discharge}(d,X)\big) = \begin{cases} \frac{\mu X}{W}D(d) & \text{for } X \in I_S, I_{ST} \\ \frac{X}{W}D(d) & \text{for } X \notin I_S, I_{ST} \end{cases} \qquad 11$$

$$W = S + S_T + E + E_T + E_a + E_{aT} + E_s + E_{sT} + I_a + I_{aT} + \mu(I_s + I_{sT}) + R_p + R_{pT} + R + R_T \qquad 12$$

As with initiations above, we denote the number of patients discharged on day *d* in compartment *X* by Equation 13, which is equal to the integral of all discharges in that compartment across the whole of the day. The total discharges on day *d* across all compartments *U* is equal to *D*(*d*) (Equation 14).

$$\textbf{Discharge}(d, X) = \int_{t=d}^{t=d+1} \textbf{Discharge}(t, X) dt \quad (13)$$

$$\sum_{X \in U} \textbf{Discharge}(d, X) = D(d) \quad (14)$$

##### *Testing model*

The expectation of the number of tests to occur in a compartment *X* on day *d* is given as follows, with *T*(*d*) referring to the number of tests occurring on day *d* in the data (Equation15).

The untested symptomatic patients are tested as a priority and so the number of tests they receive is determined as the minimum of their size ($I_s$) and the number of tests available (*T*(*d*)), so the expected number of these (in Equation 15 for $I_s$) is the same as their total (Equation 18).

The remaining tests are distributed randomly throughout the remaining compartments. Expectations are shown in Equation 15, derived from the number of tests left over after first symptomatic tests, the size of the compartment, the parameter $\varphi$ for the already tested compartments, and a denominator *M* (Equation 16) which represents the total testable population (including the adjustment for retesting).

$$E(\textbf{Test}(d, X)) = \begin{cases} \min(T(d), I_S) & \text{for } X \in I_S \\ \min(T(d) - I_S, 0)\dfrac{X}{M} & \text{for } X \in S, E, E_a, E_s, I_a, R_p, R \\ \min(T(d) - I_S, 0)\dfrac{\varphi X}{M} & \text{for } X \in S_T, E_T, E_{aT}, E_{sT}, I_{aT}, I_{sT}, R_{pT}, R_T \end{cases} \quad (15)$$

$$M = (S + E + E_a + E_s + I_a + R_p + R) + \varphi(S_T + E_T + E_{aT} + E_{sT} + I_{aT} + I_{sT} + R_{pT} + R_T) \quad (16)$$

As with initiations above, we denote the number of patients tested on day *d* in compartment *X* by Equation 17, which is equal to the integral of all tests in that compartment across the whole of the day. The total tests across all compartments other than $I_s$ is equal to the number of remaining tests (Equation 19).

$$\textbf{Test}(d, X) = \int_{t=d}^{t=d+1} \textbf{Test}(t, X) dt \quad (17)$$

$$\textbf{Test}(d, I_S) = \min(T(d), I_S) \quad (18)$$

$$\sum_{X \notin I_S} \textbf{Test}(d, X) = \min(T(d) - I_S, 0) \quad (19)$$

When simulating the observed tests on a given day using *rmeasure* in *pomp*, the number of positive and negative tests for a given day is drawn from the number of tests occurring in each compartment (Equation 15) according to the probability that a sample taken from an individual in that compartment would test positive, which is governed by specificity, *v*, for virus-free compartments, and sensitivity $z_X$ for each compartment $X$ (Equations 20 and 21). These expected distributions are also used for evaluating the likelihood of observed data using *dmeasure* in *pomp*.

$$\begin{aligned} E(\text{Positives}(d)) = {} & (\mathbf{Test}(d,S) + \mathbf{Test}(d,S_T))(1-v) + (\mathbf{Test}(d,E) + \mathbf{Test}(d,E_T))z_E \\ & + (\mathbf{Test}(d,E_a) + \mathbf{Test}(d,E_{aT}))z_{Ea} + (\mathbf{Test}(d,E_s) + \mathbf{Test}(d,E_{sT}))z_{Es} \\ & + (\mathbf{Test}(d,I_a) + \mathbf{Test}(d,I_{aT}))z_{Ia} + (\mathbf{Test}(d,I_s) + \mathbf{Test}(d,I_{sT}))z_{Is} \\ & + \left(\mathbf{Test}(d,R_p) + \mathbf{Test}(d,R_{pT})\right)z_{Rp} + (\mathbf{Test}(d,R) + \mathbf{Test}(d,R_T))(1-v) \end{aligned} \tag{20}$$

$$\begin{aligned} E(\text{Negatives}(d)) = {} & (\mathbf{Test}(d,S) + \mathbf{Test}(d,S_T))v + (\mathbf{Test}(d,E) + \mathbf{Test}(d,E_T))(1-z_E) \\ & + (\mathbf{Test}(d,E_a) + \mathbf{Test}(d,E_{aT}))(1-z_{Ea}) + (\mathbf{Test}(d,E_s) + \mathbf{Test}(d,E_{sT}))(1-z_{Es}) \\ & + (\mathbf{Test}(d,I_a) + \mathbf{Test}(d,I_{aT}))(1-z_{Ia}) + (\mathbf{Test}(d,I_s) + \mathbf{Test}(d,I_{sT}))(1-z_{Is}) \\ & + \left(\mathbf{Test}(d,R_p) + \mathbf{Test}(d,R_{pT})\right)(1-z_{Rp}) + (\mathbf{Test}(d,R) + \mathbf{Test}(d,R_T))v \end{aligned} \tag{21}$$

#### Implementation of the stochastic model

The hybrid stochastic model was implemented using *rprocess* in *pomp* using the Gillespie algorithm as the number of events on a given day is relatively small (around 15-30) given the population of less than 400 patients. The algorithm calculates a rate for each possible type of event to occur, determines the time and type of the next event accordingly, and then recalculates the rates after each event. In order to ensure that deterministic transitions (i.e. events determined by model input) occur with certainty within the framework of the Gillespie algorithm, the rate of such events was set to an arbitrarily large number, $L$=$10^6$, if further instances of that event are still to occur on day $d$. Once all required instances have occurred, the rate is set to zero.

#### Undetected epidemic dynamics

Due to repeat testing it is not possible to calculate exactly the number of infections which were detected and undetected in simulated data, but Equations 22 provide an approximation, where undetected includes both untested patients as well as false negatives.

$$\begin{aligned} &\text{Prevalence of undetected asymptomatics} = \left(E_a + I_a + E_{aT}(1-Z_{Ea}) + I_{aT}(1-Z_{Ia})\right) \\ &\text{Prevalence of undetected symptomatics} = \left(E_s + I_s + E_{sT}(1-Z_{Es}) + I_{sT}(1-Z_{Is})\right) \\ &\text{Prevalence of detected asymptomatics} = (E_{aT}Z_{Ea} + I_{aT}Z_{Ia}) \\ &\text{Prevalence of detected symptomatics} = (E_{sT}Z_{Es} + I_{sT}Z_{Is}) \end{aligned} \tag{22}$$

### B - Data and parameter estimation from data

All patients were included in the study if they were present in the hospital during the study period from day -10 to day 50 inclusive. Daily data on tests, admissions and discharges were input directly to the model from the hospital data (Figure 8). In most cases patients were recorded as present in a particular ward, and these were used to create ward-specific datasets. In case of a gap in the patient record between recorded stays in different wards the transfer was assumed to occur at the midpoint of the gap.

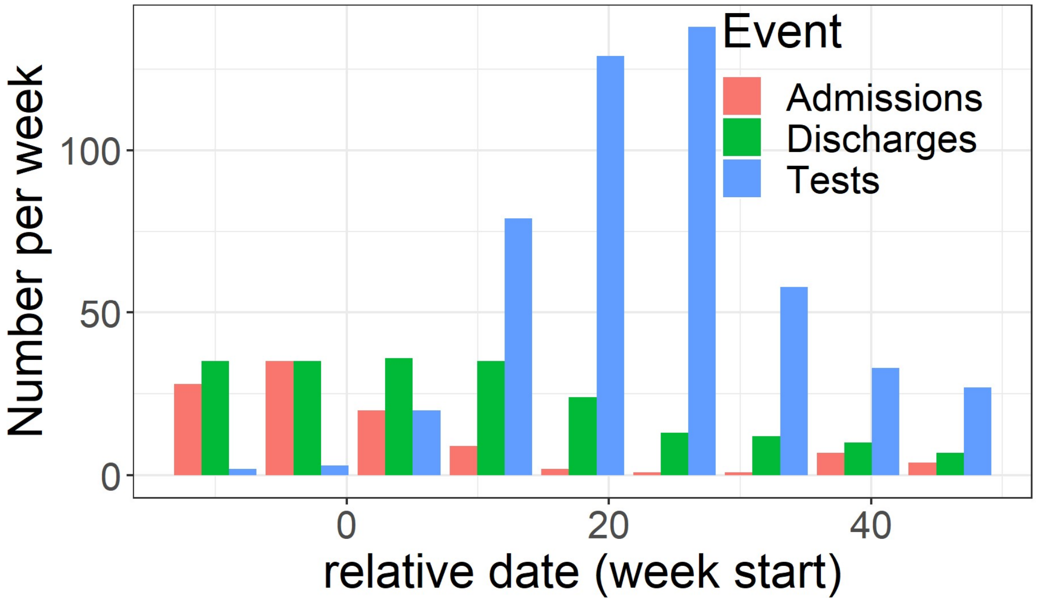


*Figure 8. Weekly aggregated numbers of admissions (red), discharges (green) and PCR tests (blue) reported over the study period. The daily disaggregated data are used in the model as A(d), D(d) and T(d) respectively.*

#### Parameters estimated directly from the longitudinal hospital data

The parameter 1/ω, the duration of the $R_p$ stage during which individuals who have recovered from infectious disease continue to frequently test positive through PCR, was selected by calculating the likelihood of each duration according to the results of repeat tests. The data used were repeat tests taken after an individual had had an earlier positive test (Figure 9, left). We assumed that the $R_p$ stage began 7 days after the first positive test (1/δ), the probability of testing positive during this stage was 30% ($Z_{Rp}$) and the probability of testing positive in the R stage which followed was 1% (1-ν) (Table 1). The likelihood reached a plateau where 1/ω = 23, so a value of 25 days was subsequently used as consistent with this result (Figure 9, right).

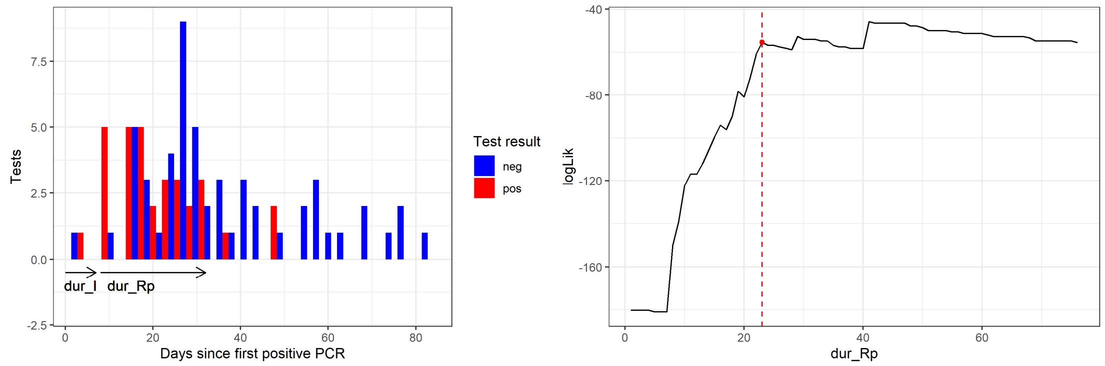


*Figure 9. Results from repeat tests taken after a first positive test (left). The duration of the full-blown infection (I) stage was 7 days according to Table 1, and the likelihood for each potential duration of the $R_p$ stage (in days, right) was calculated according to the sensitivity of the subsequent stages and the number of tests from each.*

The parameter φ, the relative rate of retesting, has been estimated crudely from the data by counting the number of repeat tests i.e. on individuals who have been tested again without having developed symptoms (211) divided by the sum of the number of first tests (314) plus the number who were retested upon developing symptoms (34), giving a retesting rate of φ = 60%. A bootstrapping analysis was conducted on the dataset to estimate CIs of 50 and 70%.

#### Likelihood calculation

The infection model was linked to observed data (number of positive and negative tests per day) using the framework of a partially observed Markov process (POMP) in which the modified SEIR model governs the underlying infection dynamics, and each day the observation process provides a likelihood of observing the data given the internal state.

The likelihood for the observation of a number of negative and positive tests on a particular day *d* is given in Equation 23. The set of parameters is represented by θ, and normally only $\beta$ and $t_{init}$ would vary within the single phase model, while in the two-phase model $\beta_1$, $\beta_2$, and $t_{init}$ are estimated. The expected numbers of positives and negatives according to the model and θ are given by Equations 20 and 21. The total likelihood is the product of the likelihood values across all time points *d*.

$$\text{Likelihood}_d = p(\text{testing negative}(d)|\text{model}, \theta)^{\text{Negatives}(d)} p(\text{testing positive}(d)|\text{model}, \theta)^{\text{Positives}(d)}$$
$$= \left(\frac{E(\text{negatives}(d))}{\left(E(\text{negatives}(d)) + E(\text{positives}(d))\right)}\right)^{\text{Negatives}(d)} \left(\frac{E(\text{positives}(d))}{\left(E(\text{negatives}(d)) + E(\text{positives}(d))\right)}\right)^{\text{Positives}(d)} \quad (23)$$

#### Parameter estimation through stochastic model fitting

The inference of parameters (transmission rates $\beta$, or $\beta_1$ and $\beta_2$ as well as $t_{init}$ or $E_{init}$) was conducted according to the methodology proposed by King et al. [20]. The first step was an initial search for the values of all parameters to be estimated, using 500 iterations of 500 particles, and a cooling fraction of 50% every 50 steps. This was repeated 10 times for each of 1000 different starting points of the parameters to be estimated.

Subsequently a likelihood profile was estimated for $\beta$ (in the one-phase model) or $\beta_1$ (in the two-phase model) by repeating the analysis above, but using starting points for the parameter to be profiled across its relevant range (e.g. 0.1 to 10 in steps of 0.1), and holding this parameter constant while estimating the other(s) using the same inference methodology.

In all iterative filtering analyses, transmission rates were allowed to vary during an iteration, while $t_{init}$ (or $E_{init}$) was only varied at the beginning of an iteration as an initial value parameter. During inference of the two-phase model, each $\beta$-value was only allowed to vary during the phase in which it took direct effect, meaning $\beta_1$ would only vary before $t_{inflect}$ and $\beta_2$ would only vary afterwards.

Both the initial search and likelihood profiling were conducted using 500 iterations of 500 particles in each analysis, each of which was repeated 10 times for each of 1000 different starting points of the parameters to be estimated.

The likelihood of the final parameter combination in each analysis (whether initial search or profiling) was then estimated by performing 10 repetitions of particle filtering with 100,000 particles, from which a linear average of the 10 likelihoods was taken.

#### Confidence intervals for estimated parameters

Confidence intervals for estimated parameters were established by identifying sets of parameters values with a likelihood above a threshold relative to the highest likelihood for each analysis. The threshold was the maximum value of the likelihood minus half of the 95% quantile of the χ-square distribution with degrees of freedom corresponding to the number of parameters to be estimated, typically two for the one-phase model ($\beta$ and $t_{init}$) and three for the two-phase model ($\beta_1$, $\beta_2$ and $t_{init}$).

#### Model inference validation

We conducted validation using synthetic data in order to test the effectiveness of the model and statistical inference to recover known values of the parameters. Values of the parameters of interest were varied simultaneously, while all other parameters were fixed as depicted in Table 1.

Datasets were generated using *rmeasure* in *pomp* for each set of known parameter values over a three month observation period (day -39 to day 50). Multiple (n=10) datasets were generated for each ward, or the whole hospital, and each set of known parameter values. Iterative filtering to estimate the relevant parameters was then conducted on each dataset. The one-phase model was validated using data at the scale of both the whole hospital and individual wards, while the two-phase model was validated only at the whole-hospital level.

Unlike in the analysis on the true data, the value of $\beta$ or $\beta_1$ was only estimated by iterative filtering directly, without the systematic likelihood profile for a range of values. For each parameter search, 500 iterations of 500 particles were used. The known values could then be compared with median values and ranges of the estimates.

In order to systematically identify wards with sufficient power to be analysed using our inference methodology, the resulting estimated values were compared with true values. For each dataset, a deviation was estimated as the ratio of the estimated to the true value. If the median value of this deviation across all analyses for the ward was less than or equal to 1.15, it was considered that the ward had sufficient power to be analysed.

### C - Additional symbols

*Table 4. Additional parameter values, state variables and their descriptions, not including those described in Table 1.*

| Symbol | Description |
|---|---|
| λ(t) | Force of infection at time *t* |
| $\kappa_2$, $\kappa_3$ | Relative rates of progression to full infection (Ea to Ia) and recovery in asymptomatic pathway, relative to symptomatic pathway (assumed 1) |
| W | The total rate adjusted number of dischargeable individuals across all compartments at a given time |
| M | The total rate adjusted number of testable individuals across all compartments including those eligible for retesting, but excluding untested $I_s$, at a given time |
| N | The total population size at a given time |
| U | The universal set of compartments |
| A(d) | The number of admissions in the data on day *d* |
| D(d) | The number of discharges in the data on day *d* |
| T(d) | The number of tests in the data on day *d* |
| C(d) | The number of infections initiated (moved from *S* to *E* or $S_T$ to $E_T$ on day *d*) |

| | |
|---|---|
| Admission(t) | Admissions occurring into compartment *S* at time *t* |
| Initiation(X,t) | Infection initiations occurring from compartment *X* at time *t* |
| Discharge(X,t) | Discharges occurring from compartment *X* at time *t* |
| Test(X,t) | Tests occurring from compartment *X* at time *t* |
| $S$ | Susceptible untested at a given time |
| $E$ | Infected uninfectious untested at a given time |
| $E_a$ | Early infectious infection on asymptomatic pathway, untested at a given time |
| $E_s$ | Pre-symptomatic infectious infection on symptomatic pathway, untested at a given time |
| $I_a$ | Full-blown infection on asymptomatic pathway, untested at a given time |
| $I_s$ | Full-blown symptomatic infection, untested at a given time |
| $R_p$ | Recovered but still shedding virus, untested at a given time |
| $R$ | Recovered and no longer shedding virus, untested at a given time |
| $S_T$ | Susceptible tested at a given time |
| $E_T$ | Infected uninfectious tested at a given time |
| $E_{aT}$ | Early infectious infection on asymptomatic pathway, tested at a given time |
| $E_{sT}$ | Pre-symptomatic infectious infection on symptomatic pathway, tested at a given time |
| $I_{aT}$ | Full-blown infection on asymptomatic pathway, tested at a given time |
| $I_{sT}$ | Full-blown symptomatic infection, tested at a given time |
| $R_{pT}$ | Recovered but still shedding virus, tested at a given time |
| $R_T$ | Recovered and no longer shedding virus, tested at a given time |
