## Supplementary Results for "How well does SARS-CoV-2 spread in hospitals?"

### A - Validation of the statistical inference of model parameters

Simultaneous estimation of known parameter values was conducted for both the one- and two-phase models on data at the scale of the whole hospital. Figure 10 describes the relationship between known and estimated values of $\beta$ and $t_{init}$ for the one-phase model. The value of β was well estimated throughout the range, while the estimate of $t_{init}$ was slightly over-estimated from days -25 to -16, but not after this point.

Figure 11 describes the relationship for the two-phase model between known values of $\beta_1$, $\beta_2$ and $t_{init}$ and their estimates. The first phase transmission rate, $\beta_1$, was reliably estimated up to 1.0 with a slight overestimation after that point. The second phase transmission rate, $\beta_2$, was correctly estimated, with the exception of values wrongly estimated to be close to zero. However, in the absence of estimates close to zero in the analysis of true data, the estimates could be considered reliable. As with the one-phase model, the estimate of $t_{init}$ deviated slightly when it fell in the first weeks but was more reliable after day -15.

The ability of our framework to correctly estimate parameter values at the ward level was limited due to much smaller population sizes and specific distributions of tests. The ability to estimate values of $\beta$ and $t_{init}$ simultaneously using the one-phase model on individual wards is shown in Figure 12 and Figure 13 . Our results suggest that only data corresponding to wards A2, C0, C1, C2 and C3 provided sufficient power to be analysed through our framework, and C1 was also excluded due to the lack of visible relationship between true and estimated values.

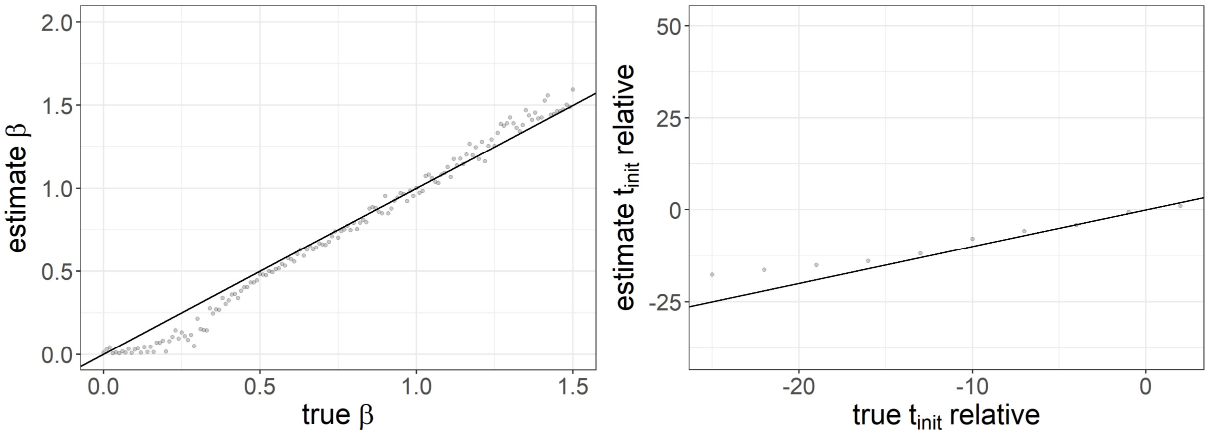


*Figure 10. Validation of simultaneous estimation of two parameters using the one-phase model on the datasets at the scale of the whole hospital. Each point represents a true value of the parameter on its x-axis, with the value on the y-axis being the median across 10 attempts to estimate the true value using particle filtering. The solid black line indicates where the true and estimated values are equal. The value of $E_{init}$ was fixed at 1.*

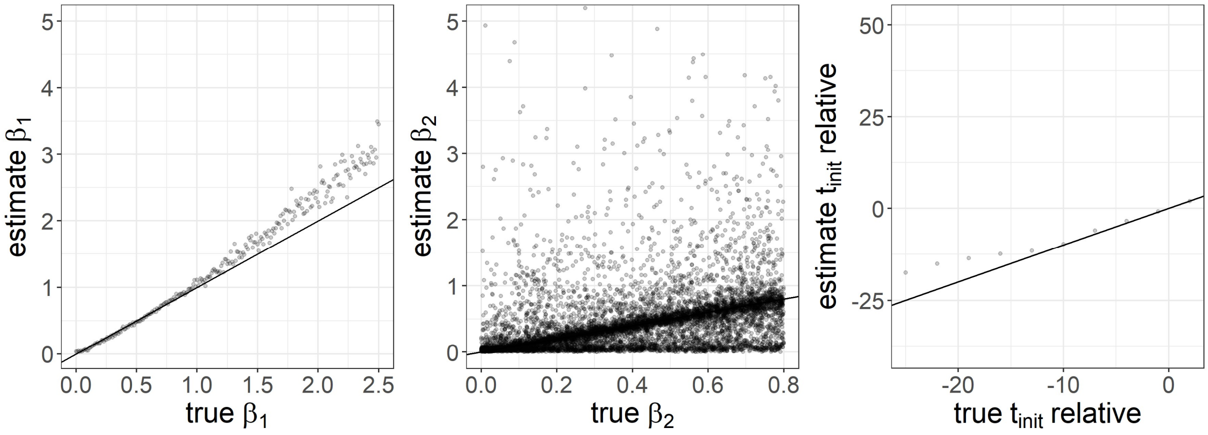


*Figure 11. Validation of simultaneous estimation of three parameters using the two-phase model on the datasets at the scale of the whole hospital. Each point represents a true value of the parameter on its x-axis, with the value on the y-axis being the median across 10 attempts to estimate the true value using particle filtering. The solid black line indicates where the true and estimated values are equal. The values of $E_{init}$ and $t_{inflect}$ were fixed at 1 and day 12, respectively.*

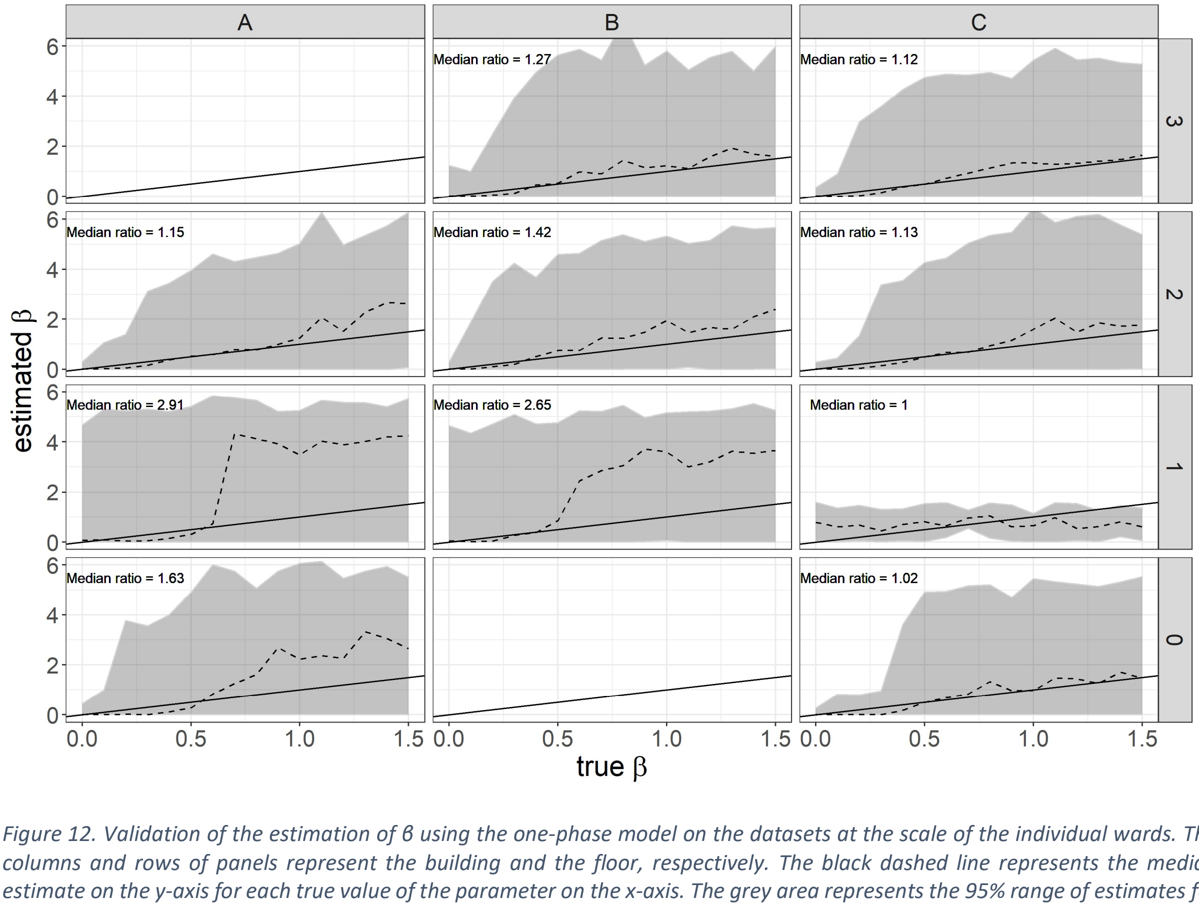


*Figure 12. Validation of the estimation of β using the one-phase model on the datasets at the scale of the individual wards. The columns and rows of panels represent the building and the floor, respectively. The black dashed line represents the median estimate on the y-axis for each true value of the parameter on the x-axis. The grey area represents the 95% range of estimates for each value of the true parameter. The solid black line indicates where the true and estimated values are equal. The value of $E_{init}$ was fixed at 1. The numerical value given in the corner is the median ratio between the estimated and true values.*

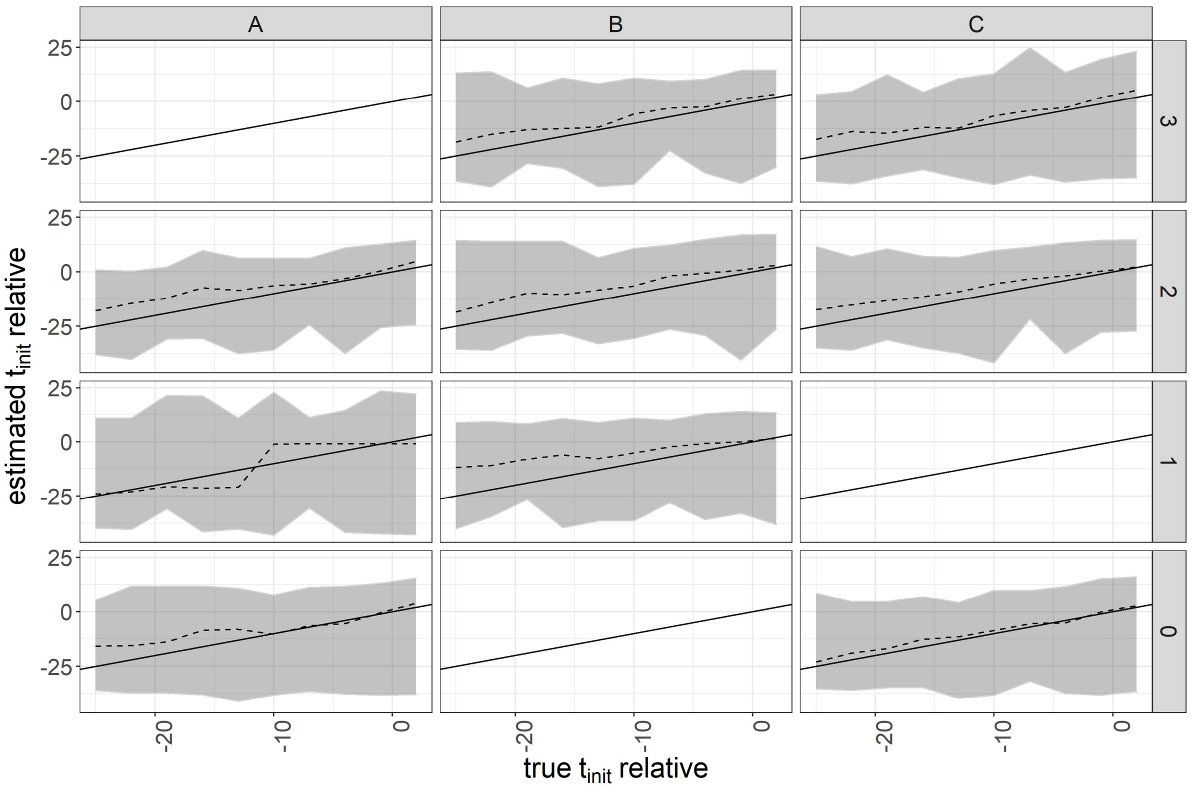


*Figure 13. Validation of the estimation of $t_{init}$ using the one-phase model on the datasets at the scale of the individual wards. The columns and rows of panels represent the building and the floor, respectively. The black dashed line represents the median estimate on the y-axis for each true value of the parameter on the x-axis. The grey area represents the 95% range of estimates for each value of the true parameter. The solid black line indicates where the true and estimated values are equal. The value of $E_{init}$ was fixed at 1.*

### B - Results of the analysis at the whole hospital level

The scatterplots in Figure 14 show the estimated likelihood profiles for $\beta$ and likelihood values for $t_{init}$ in the one-phase model with fixed values of $E_{init}$. Figure 15 shows an alternative analysis in which the values of $t_{init}$ are held constant and $E_{init}$ is allowed to vary. Numerical results for all analyses of the one-phase model are shown in Table 5, with those for $E_{init}$ = 1 also in Table 2.

The scatterplots in Figure 16 show the final estimated likelihood for each combination of parameters in the two-phase model, with fixed values of $E_{init}$. An alternative analysis, in which $t_{init}$ was fixed and $E_{init}$ was estimated (Figure 17) provided similar transmission rates. Numerical results for all analyses of the two-phase model with $t_{inflect}$ = day 12 are shown in Table 7, with those for $E_{init}$ = 1 also in Table 2. Analyses exploring the effect of changing $t_{inflect}$ are shown in Table 6.

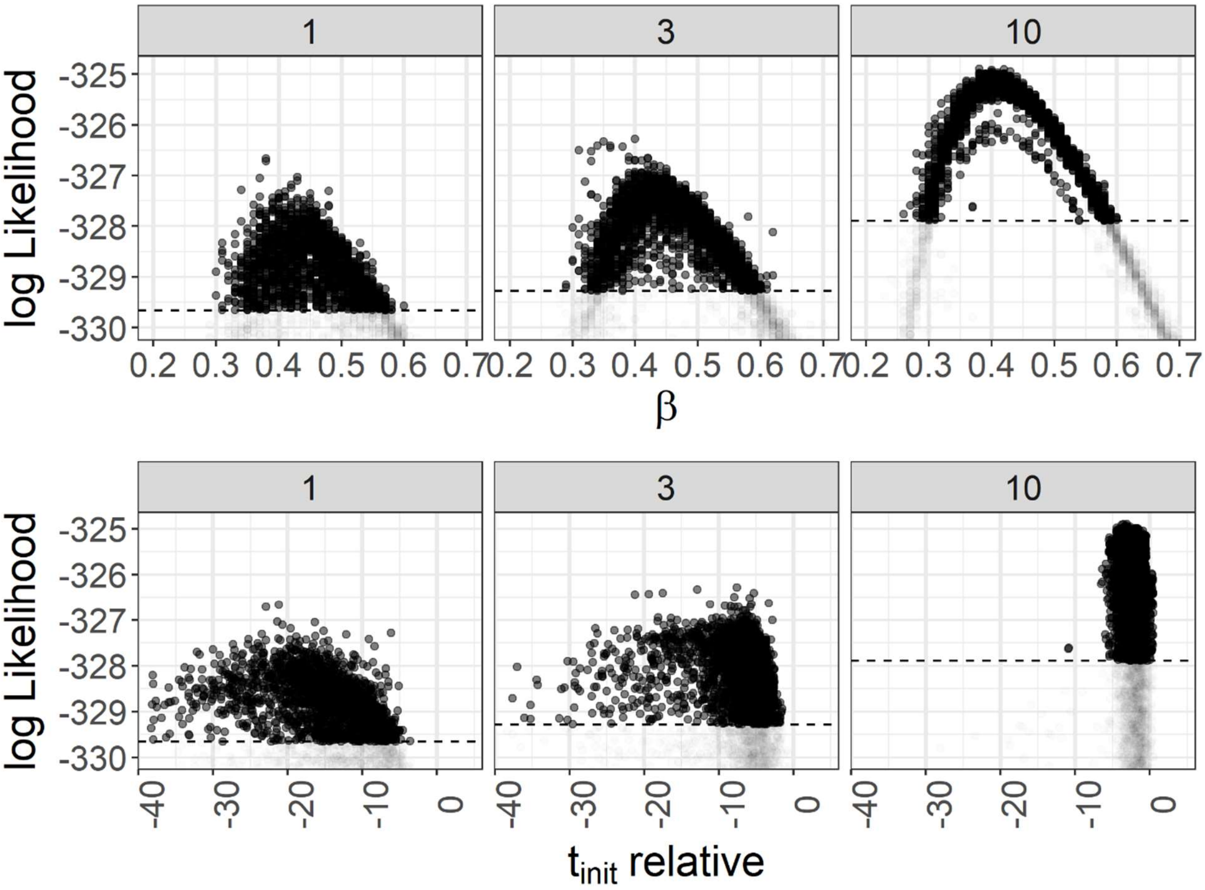


*Figure 14. Likelihood profile for β and* $t_{init}$ *estimated by iterative filtering for fixed values of* $E_{init}$ *(indicated by the grey bars at the top of each plot) in a model with only a single transmission rate β. The dashed line indicates the 95% confidence limit for likelihood values relative to the point of highest likelihood.*

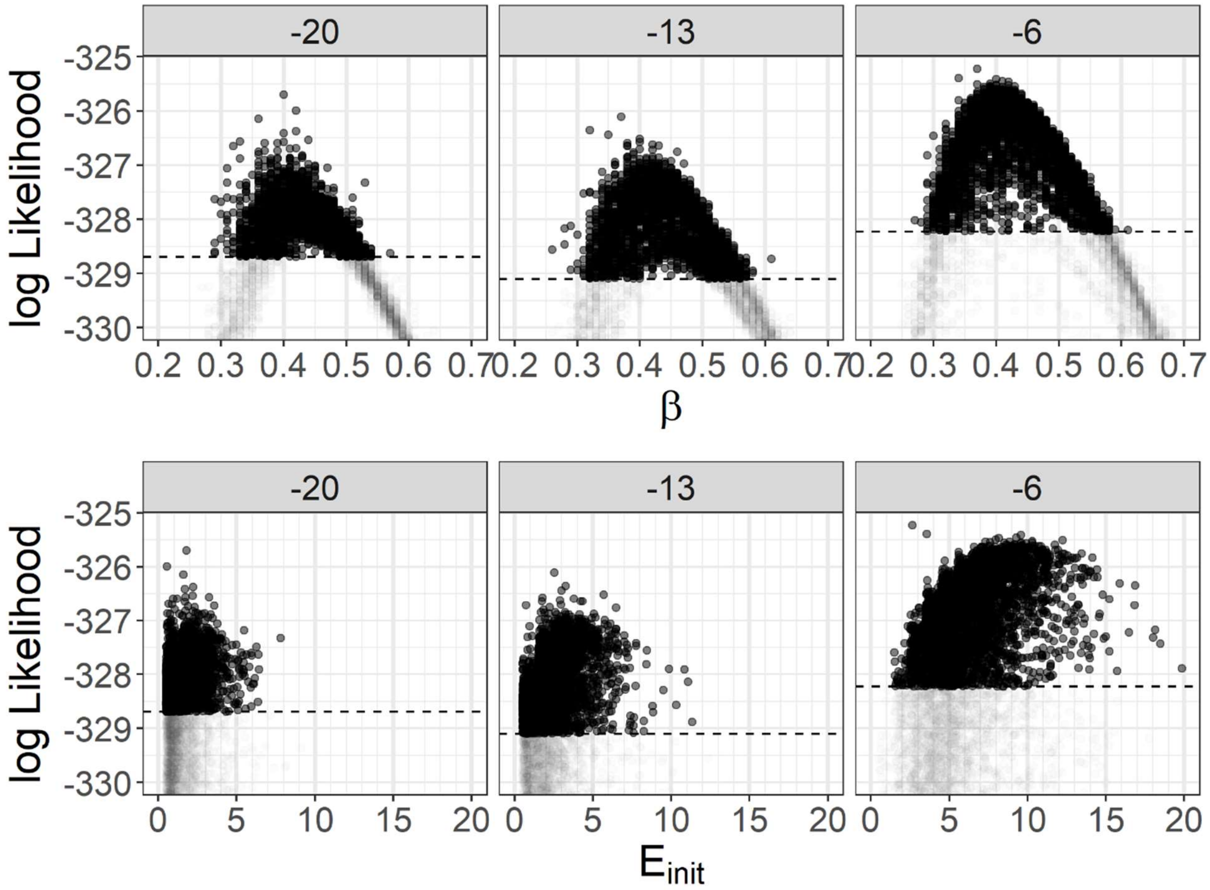


*Figure 15. Likelihood profile for β and $E_{init}$ estimated by iterative filtering for fixed values of $t_{init}$ (indicated by the grey bars at the top of each plot) in a model with only a single transmission rate β. The dashed line indicates the 95% confidence limit for likelihood values relative to the point of highest likelihood.*

*Table 5. Best estimates and the ranges for β in a single transmission rate model, with corresponding $R_0$ values, along with fixed values (bold) or ranges for $E_{init}$ or $t_{init}$. The $R_0$ values are calculated using Equation 4. Values for $t_{init}$ are relative to the start of the study data.*

| Estimate | *β* | $R_0$ | $E_{init}$ | $t_{init}$ | *AIC* |
|---|---|---|---|---|---|
| **β, $t_{init}$** | 0.38 (0.30-0.60) | 2.6 (2.0- 4.1) | **1** | -22 (-39 – -4) | 657.3257 |
| | 0.40 (0.29-0.62) | 2.7 (2.0- 4.2) | **3** | -8 (-38 – -2) | 656.5639 |
| | 0.38 (0.26-0.60) | 2.6 (1.8- 4.1) | **10** | -4 (-11 – 0) | 653.7993 |
| **β, $E_{init}$** | 0.37 (0.27-0.61) | 2.5 (1.8- 4.1) | 2.7 (1.5-19.9) | **-6** | 654.4575 |
| | 0.37 (0.26-0.61) | 2.5 (1.8- 4.1) | 2.5 (0.5-11.3) | **-13** | 656.2111 |
| | 0.40 (0.29-0.57) | 2.7 (2.0- 3.9) | 1.8 (0.5- 7.8) | **-20** | 655.3993 |

*Table 6. Effect of changing $t_{inflect}$ on parameter values. Best estimates and their ranges for $\beta_1$, $\beta_2$, their corresponding $R_0$ values, and $t_{init}$. The risk ratio is calculated for each point estimate as $\beta_1/\beta_2$. The $R_0$ values are calculated using Equation 4, substituting the corresponding $\beta$ value. The combined $R_0$ is an average $R_0$ in each phase weighted by phase duration (Equation 5).*

| $t_{inflect}$ | $\beta_1$ | $\beta_2$ | $R_{0\ before}$ | $R_{0\ after}$ | $R_{0\ combined}$ | Intervention efficacy | $t_{init}$ | AIC |
|---|---|---|---|---|---|---|---|---|
| **1** | 3.44 (0.29-49.50) | 0.36 (0.23-0.57) | 23.38 (2.00-336.67) | 2.47 (1.60-3.88) | 13.59 (2.41-180.07) | 0.89 (-0.47-0.99) | -4 (-37-1) | 654.27 |
| **6** | 2.68 (0.99-49.50) | 0.29 (0.17-0.40) | 18.21 (6.71-336.67) | 1.98 (1.16-2.70) | 10.61 (4.50-179.87) | 0.89 ( 0.64-1.00) | -2 (-19-6) | 639.97 |
| **8** | 2.24 (0.99-12.10) | 0.25 (0.16-0.34) | 15.22 (6.73- 82.30) | 1.68 (1.07-2.35) | 8.87 (4.39- 44.43) | 0.89 ( 0.68-0.98) | -2 (-20-3) | 633.56 |
| **10** | 1.62 (0.91- 4.00) | 0.22 (0.14-0.32) | 11.01 (6.18- 27.21) | 1.49 (0.96-2.14) | 6.55 (4.14- 15.12) | 0.86 ( 0.67-0.95) | -3 (-22--1) | 629.89 |
| **12** | 1.28 (0.76- 2.00) | 0.19 (0.10-0.30) | 8.72 (5.14- 13.60) | 1.33 (0.68-2.04) | 5.26 (3.38- 7.94) | 0.85 ( 0.66-0.94) | -4 (-24--1) | 628.85 |
| **14** | 1.02 (0.70- 1.50) | 0.17 (0.08-0.26) | 6.91 (4.73- 10.20) | 1.15 (0.56-1.77) | 4.22 (3.06- 6.09) | 0.83 ( 0.63-0.94) | -5 (-27--2) | 630.19 |
| **16** | 0.84 (0.54- 1.08) | 0.16 (0.07-0.26) | 5.69 (3.65- 7.31) | 1.07 (0.48-1.78) | 3.53 (2.61- 4.29) | 0.81 ( 0.61-0.93) | -5 (-28--2) | 634.83 |

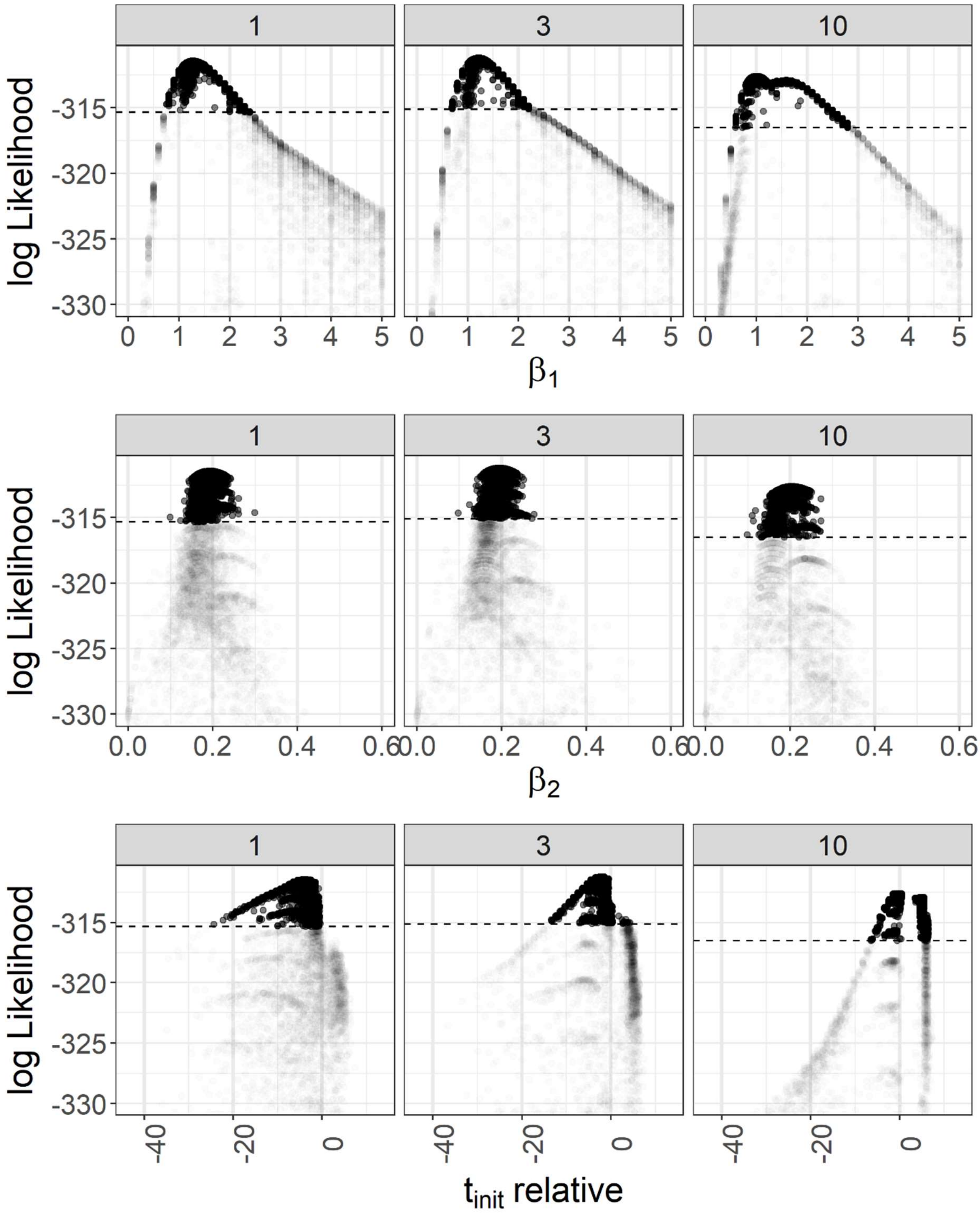


*Figure 16. Likelihood profile for $\beta_1$, $\beta_1$ and* $t_{init}$ *estimated by iterative filtering for fixed values of $E_{init}$, indicated by the grey bars at the top of each plot. The dashed line indicates the 95% confidence limit for likelihood values relative to the point of highest likelihood.*

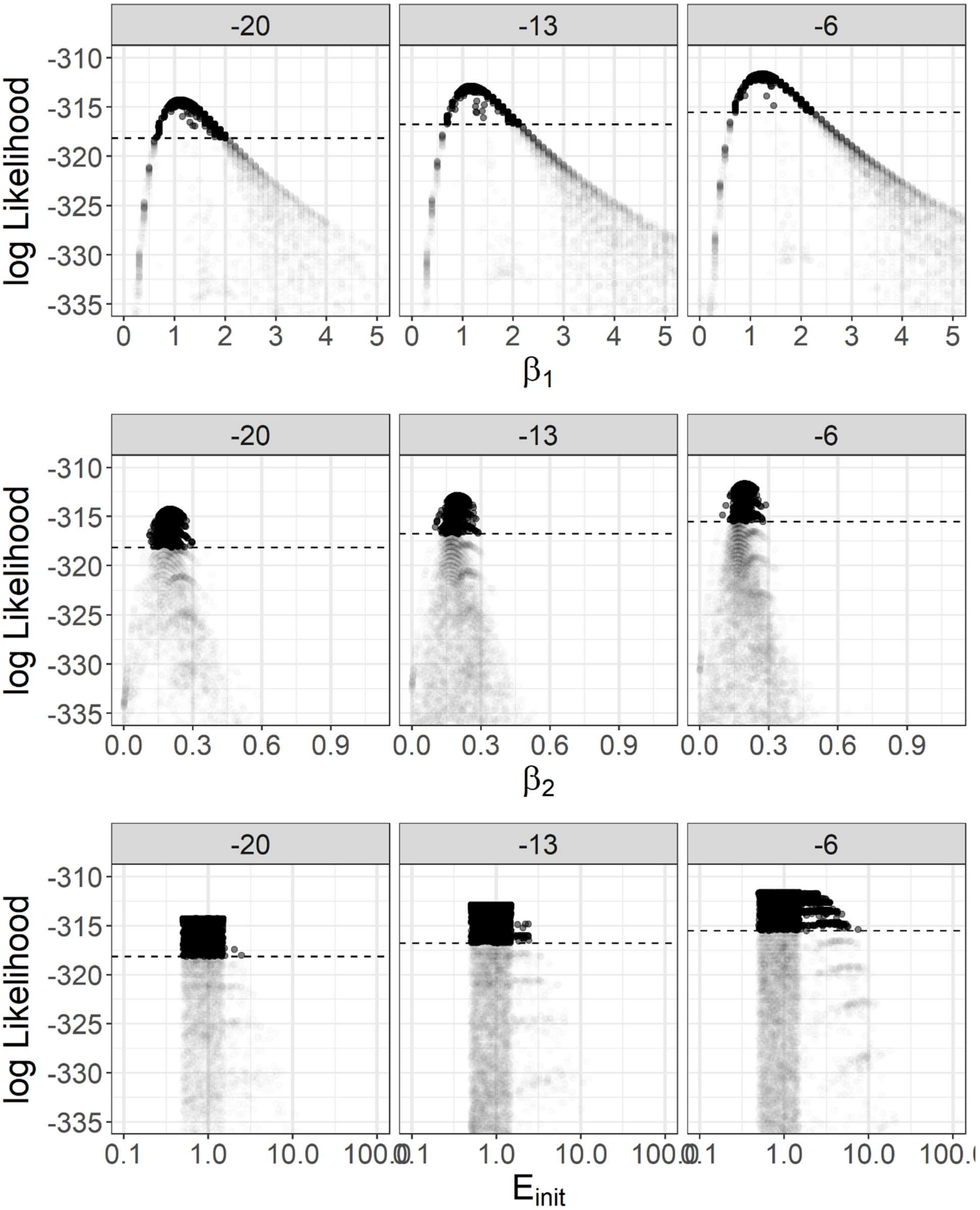


*Figure 17. Likelihood profile for $\beta_1$, $\beta_1$* and *$E_{init}$ estimated by iterative filtering for fixed values of $t_{init}$, indicated by the grey bars at the top of each plot. The dashed line indicates the 95% confidence limit for likelihood values relative to the point of highest likelihood.*

*Table 7. Best estimates and their ranges for $\beta_1$, $\beta_2$, their corresponding $R_0$ values, along with fixed values (bold) or ranges for $E_{init}$ or $t_{init}$. The $R_0$ values were calculated using Equation 4, substituting the corresponding $\beta$ value.*

| Estimate | $E_{init}$ | $\beta_1$ | $\beta_2$ | $R_0$ before | $R_0$ after | $t_{init}$ | AIC |
|---|---|---|---|---|---|---|---|
| $\beta_1$, $\beta_2$, $t_{init}$ | **1** | 1.28 (0.76-2.40) | 0.19 (0.10-0.30) | 8.72 (5.14-16.32) | 1.33 (0.68-2.04) | -4 (-24 – 0) | 628.85 |
| | **3** | 1.23 (0.68-2.20) | 0.19 (0.10-0.28) | 8.37 (4.65-14.96) | 1.31 (0.66-1.89) | -2 (-13 – 4) | 628.4 |
| | **10** | 1.03 (0.59-2.80) | 0.20 (0.10-0.27) | 7.03 (4.00-19.04) | 1.39 (0.67-1.85) | 0 (-6 – 6) | 631.21 |
| $\beta_1$, $\beta_2$, $E_{init}$ | 0.70 (0.50-2.49) | 1.10 (0.63-2.00) | 0.20 (0.11-0.30) | 7.48 (4.26-13.60) | 1.39 (0.76-2.02) | **-6** | 634.51 |
| | 0.63 (0.50-2.43) | 1.14 (0.69-2.10) | 0.20 (0.10-0.29) | 7.78 (4.68-14.28) | 1.36 (0.69-1.96) | **-13** | 631.74 |
| | 1.12 (0.50-7.47) | 1.24 (0.70-2.20) | 0.19 (0.10-0.29) | 8.42 (4.73-14.96) | 1.32 (0.67-1.95) | **-20** | 629.24 |

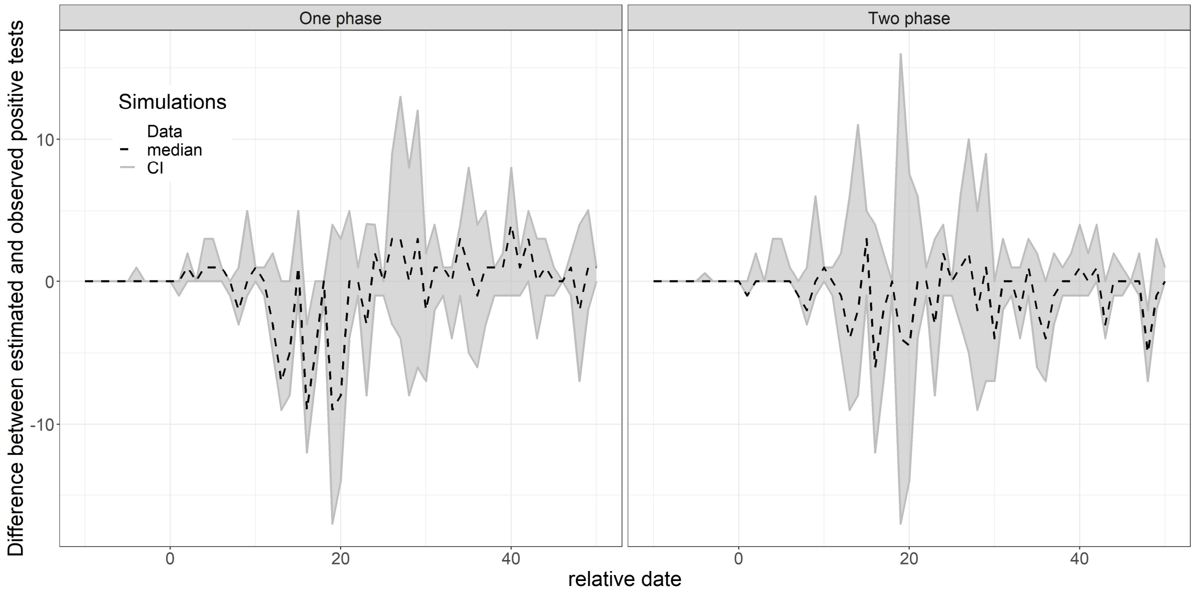


*Figure 18. Comparison between the one- and two-phase models as to the proximity of their median simulated numbers of positives tests to the true values, across the whole hospital. This is equivalent to Figure 3 but in which the value of the observed data has been subtracted at each date.*

### C - Results of the analysis at the individual ward level

Figure 19 shows the estimates of the $\beta$ and $t_{init}$ for each ward which passed model validation using the one-phase model. A comparison of results from each ward is shown in Table 8, which demonstrates that the AIC for the one-phase model is lower or equal for three out of four wards.

*Table 8. Best estimates and their ranges $\beta_1$, $\beta_2$, $R_0$ in each phase and combined and $t_{init}$ for the two-phase model across each ward. The values of $E_{init}$ and $t_{inflect}$ were fixed at 1 and day 11 respectively. In many instances, the upper bound of the CI for $\beta_1$, and in some the most likely value of $\beta_1$ as well, could not be estimated due to a flat likelihood surface, in which case the value is given as N.E.=not estimated. The risk ratio is calculated for each point estimate as $\beta_1/\beta_2$. The $R_0$ values were calculated using Equations 4 and 5.*

| | Two-phase | One-phase |
|---|---|---|

| Ward | β1 | β2 | Risk ratio | $R_{0\ combined}$ | $t_{init}$ | AIC | AIC |
|---|---|---|---|---|---|---|---|
| A2 | 2.16 (0.30-N.E.) | 0.70 (0.31-4.42) | 0.33 (0.04- 11.39) | 10.41 (4.77-49.01) | 4 (-20 – 7) | 139.4 | 138.25 |
| C0 | N.E. | 0.35 (0.26-4.89) | 0.04 (0.03- 7.20) | 39.44 (1.75-50.23) | 10 (-38 – 11) | 89.91 | 91.59 |
| C2 | N.E. | 0.00 (0.00-0.08) | 0.00 (0.00- 0.06) | 37.25 (1.16-38.52) | -14 (-33 – -12) | 57.62 | 57.92 |
| C3 | 6.50 (0.00-N.E.) | 0.41 (0.23-0.64) | 0.06 (0.03- 18353.26) | 26.43 (1.03-39.82) | 20 (-26 – 21) | 47.25 | 45.25 |

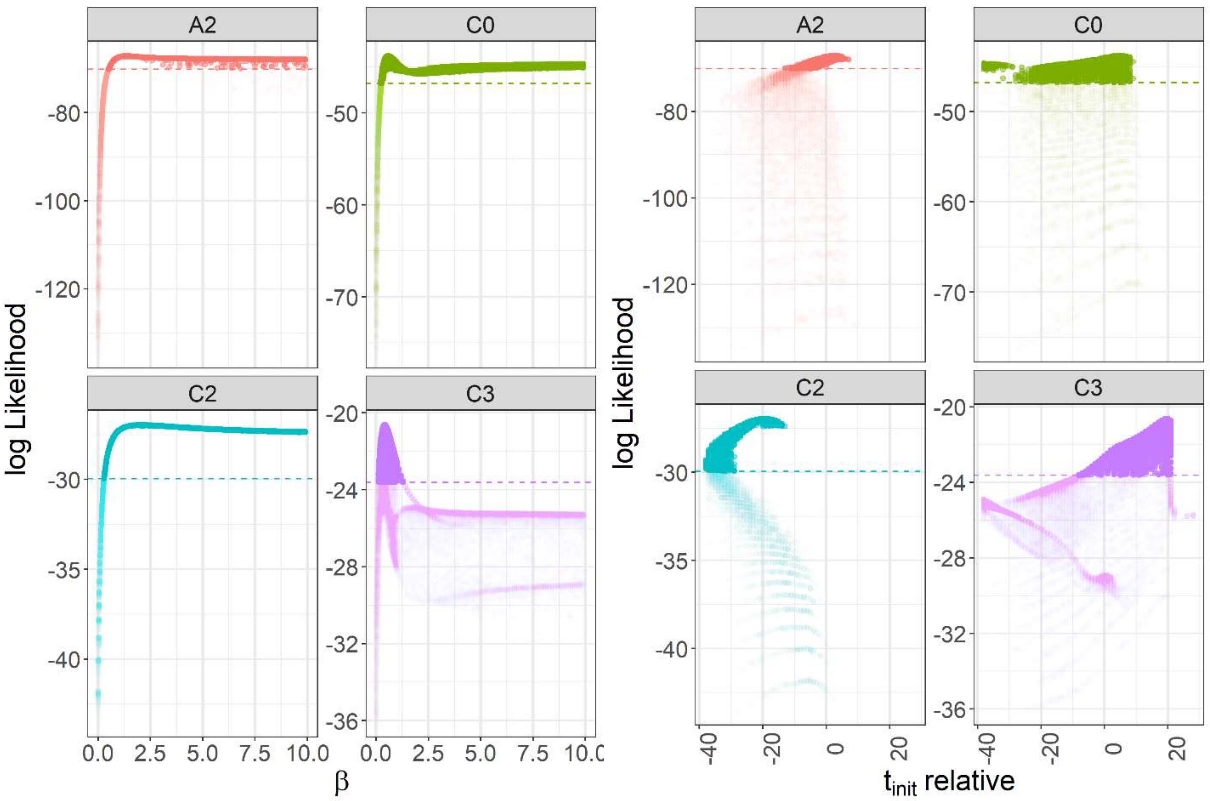


*Figure 19. Likelihood profiles of β (left four panels) and $t_{init}$ (right four panels) for each ward (indicated by the grey bars at the top of each plot) estimated by iterative filtering for $E_{init}$=1. The dashed line indicates the 95% confidence limit for likelihood values relative to the point of highest likelihood.*

### D - Sensitivity analysis

Best estimates and CI for $\beta_1$, $\beta_2$ and $t_{init}$ are shown in Figure 20. Many parameters affect the upper ranges of $\beta_1$, most markedly $\varepsilon$ which also affects the best estimate. However, the relative effects on $\beta_2$ are much greater, with $\delta$ and the sensitivity parameters of form $Z_x$ having a large effect on both the mean estimate and range. An early inflection point, $t_{inflect}$, serves to suggest the possibility of an earlier epidemic initiation point, $t_{init}$, but the biggest effect on $t_{init}$ comes from perturbing the number of index cases, $E_{init}$, with a larger number of indices pointing to a later introduction.

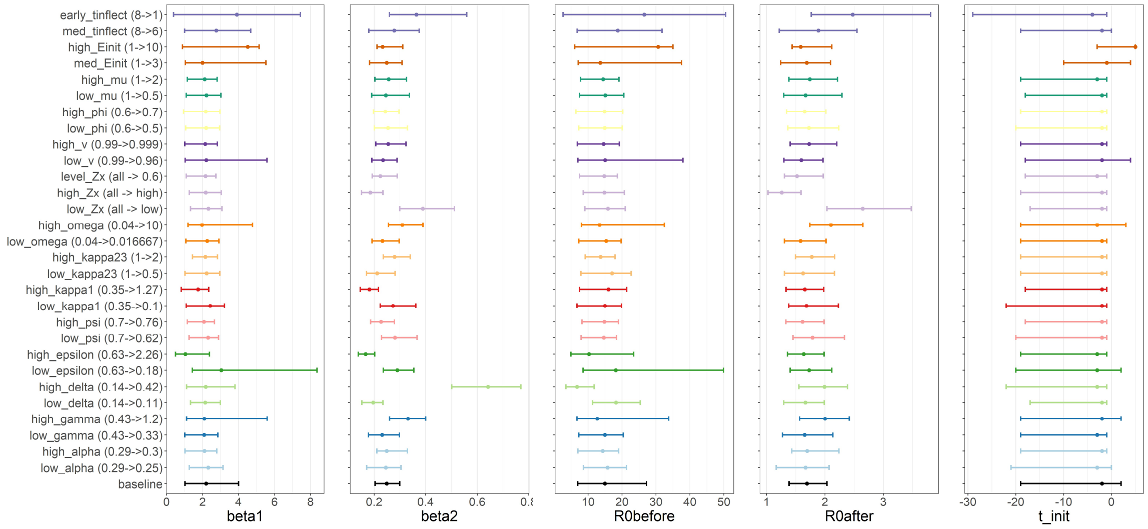


*Figure 20. Results of sensitivity analysis, showing the estimation of $\beta_1$, $\beta_2$ and their corresponding $R_0$ values $R_{0before}$ and $R_{0after}$, and $t_{init}$ under parameter values perturbed according to their uncertainty ranges, as in Table 1, with all other parameters held at baseline values. The name of the scenario is given on the left, with the change in parameter magnitude indicated. The corresponding dot shows the values of the parameter values which had the highest likelihood, while the error bars show the 95% CI. In the scenario modifying $Z_x$, all $Z_x$ parameters were modified simultaneously, to their lower or upper bounds, or to 0.6. In the scenario "kappa23" both $\kappa_2$ and $\kappa_3$, the relative rates of progression from stages* $E_a$ and $I_a$, respectively, compared to the equivalent symptomatic stage, *were modified by the same factor.*
